# Associations of Sputum Biomarkers with Clinical Outcomes in People with Cystic Fibrosis

**DOI:** 10.1101/2022.05.25.22275540

**Authors:** Theodore G Liou, Natalia Argel, Fadi Asfour, Perry S Brown, Barbara A Chatfield, David R Cox, Cori L Daines, Dixie Durham, Jessica A Francis, Barbara Glover, My Helms, Theresa Heynekamp, John R Hoidal, Judy L Jensen, Christiana Kartsonaki, Ruth Keogh, Carol M Kopecky, Noah Lechtzin, Yanping Li, Jerimiah Lysinger, Osmara Molina, Craig Nakamura, Kristyn A Packer, Robert Paine, Katie R Poch, Alexandra L Quittner, Peggy Radford, Abby J Redway, Scott D Sagel, Rhonda D Szczesniak, Shawna Sprandel, Jennifer L Taylor-Cousar, Jane B Vroom, Ryan Yoshikawa, John P Clancy, J Stuart Elborn, Kenneth N Olivier, Frederick R Adler

## Abstract

**Background:** Airway inflammation promotes bronchiectasis and lung injury in cystic fibrosis (CF). Amplification of inflammation underlies pulmonary exacerbations of disease. We asked whether sputum inflammatory biomarkers provide explanatory information on pulmonary exacerbations.

**Patients and Methods:** We collected sputum from randomly chosen stable adolescents and adults and prospectively observed time to next exacerbation, our primary outcome. We evaluated relationships between potential biomarkers of inflammation, clinical characteristics and outcomes and assessed clinical variables as potential confounders or mediators of explanatory models. We assessed associations between the markers and time to next exacerbation using proportional hazard models adjusting for confounders.

**Results:** We enrolled 114 patients, collected data on clinical variables [December 8, 2014 to January 16, 2016; 46% male, mean age 28 years (SD 12), mean percent predicted forced expiratory volume in 1 s (FEV_1_%) 70 (SD 22)] and measured 24 inflammatory markers. Half of the inflammatory markers were plausibly associated with time to next exacerbation. Age and sex were confounders while we found that FEV_1_% was a mediator.

Three potential biomarkers of RAGE axis inflammation were associated with time to next exacerbation while six potential neutrophil-associated biomarkers indicate associations between protease activity or reactive oxygen species with time to next exacerbation.

**Conclusion:** Pulmonary exacerbation biomarkers are part of the RAGE proinflammatory axis or reflect neutrophil activity, specifically implicating protease and oxidative stress injury. Further investigations or development of novel anti-inflammatory agents should consider RAGE axis, protease and oxidant stress antagonists.

**Tweetable abstract:** Sputum from 114 randomly chosen people with CF show RAGE axis inflammation, protease and oxidative stress injury are associated with time to next pulmonary exacerbation and may be targets for bench or factorial design interventional studies. (242 characters)

## Introduction

Airway inflammation erodes lung health in cystic fibrosis (CF). Infecting organisms initiate and amplify inflammation in infants with CF (1). Infections persist, interact (2) and lead to airway obstruction, mucus impaction, antibiotic resistant microbial biofilms and bronchiectasis. Consequences include strenuous treatment burdens, reduced quality of life, hospitalizations for pulmonary exacerbations and early mortality (3).

Nine variables summarize clinical CF and durably and successfully predict survival (4, 5) but do not explain inflammatory pathways underlying clinical disease and outcomes. Seeking explanations quantitatively linking inflammation to clinical outcomes, we previously found that high mobility group box 1 (HMGB1) adjusted by number of prior pulmonary exacerbations was associated with time to next exacerbation in a proportional hazards model (6). Other biomarkers may help identify and explain inflammatory pathway roles in CF. In particular, calprotectin (a heterodimer of S100A8 and S100A9), neutrophil elastase (NE) were associated with subsequent pulmonary exacerbations (7–9) in single center studies. None of the study cohorts fully represented people with CF, and no study considered confounding (10) or mediation (11) of inflammatory relationships by clinical covariables.

To potentially validate calprotectin, NE or HMGB1, or identify other biomarkers as explanatory for clinical CF outcomes, we performed a prospective observational multicenter study incorporating randomized selection to minimize observer bias and maximize generalizability (12). We collected sputum during clinical stability as the least invasive sample closest to sources of persistent airway inflammation and evaluated clinical covariables for confounding and mediation of biomarker effects leading to a pulmonary exacerbation. This strategy does not seek a prediction model (13), but rather searches for insights into pathways underlying pulmonary exacerbations in CF, seeking specific targets for further investigation and potentially novel interventions.

## Methods

### Study Design and Patients

We previously published a detailed study design (12). Briefly, we randomly selected patients with CF 12 years and older from accredited care centers for a prospective observational study of sputum biomarkers of airway inflammation. Our primary outcome was time to next pulmonary exacerbation defined as the number of days between enrollment and first hospitalization for acute treatment of at least one symptom and one objective sign of exacerbation of CF (6).

After institutional reviews (Supplemental Table 1), we obtained written informed consent from adults and assent from adolescents with parental consent before enrolling clinically stable participants at outpatient visits from December 8, 2014 through January 16, 2016. We collected, processed, aliquoted, froze expectorated sputum samples within 4 hours of collection (mean 54 minutes, SD = 98 minutes, max = 225 minutes on ice) and annotated with clinical information (12). We measured forced expiratory volume in 1 s (FEV_1_) according to American Thoracic Society Guidelines (14) and estimated percent predicted FEV_1_ (FEV_1_%) using third National Health and Nutrition Examination Study (NHANES III) equations (15). Prior year exacerbations counts were based on hospitalization dates within one year prior to enrollments. We calcuated weight-for-age *z*-score and 5-year prognostic scores, with higher scores indicating longer predicted survival (4).

### Biomarker Measurements

The Pediatric Clinical Translational Research Center Core Laboratory at Colorado Children’s Medical Center measured NE activity spectrofluorometrically (7). The University of Utah CF Center lab measured Calprotectin, C-reactive protein (CRP), and HMGB1 by enzyme linked immunosorbent assays (ELISA) using commercially available antibodies (Supplemental Table 2). We shipped samples on dry ice (Fedex, Memphis, TN, USA) to R&D Systems’ Biomarker Testing Service to assay remaining cytokines using their Human Magnetic Luminex Screening Assay Premixed Multi-Analyte Kits (Cat # LXSAHM-16 and LXSAHM-04). We used quantile plots to visually ascertain (16) upper and lower limits of detection. Log-transformed concentrations above or below the limits of detection were assigned values 0.1% above or below the limits of detection, respectively, to retain the partial information.

### Statistical Analysis

We compared clinical measurements that predict CF survival (4, 5) between participants with at least one pulmonary exacerbation in the year following enrollment and those without exacerbations. We obtained Kaplan-Meier (17) estimates of the distribution of time to next pulmonary exacerbation stratified by clinical variables, followed by evaluations using logistic regression (18) and proportional hazards models (19). For logistic regression, we used the occurrence of the next exacerbation during study follow up as the outcome with clinical variables as the inputs, and for proportional hazards models, we used time to next exacerbation as the outcome with the clinical variables as explanatory variables.

We evaluated collinearity among biomarker measurements adjusted for detection limits with Pearson Product Moment Correlations (20). We considered *p <* 0.01 after Bonferroni correction as statistically significant.

Age, sex and prior pulmonary exacerbations may confound time-to-next pulmonary exacerbation but by definition cannot be mediators thus should be included to adjust models (10). To help assess whether other clinical variables were confounding, mediating or both (11) for associations of biochemical markers with time to next pulmonary exacerbation, we fitted univariable linear regression models of age, weight-for age *z*-score, diabetes and FEV_1_% as outcomes with each potential biomarker as explanatory variable. We fitted quasi-Poisson models of prior pulmonary exacerbations with each potential biomarker. We tested each potential biomarker as an adjustment to each of the remaining potentially mediating clinical variables (weight-for-age *z*-score, diabetes and FEV_1_%) in proportional hazards models for time to next pulmonary exacerbation. We excluded variables that substantially reduced biomarker effect sizes and significance, indicating a mediation effect.

We fitted a regression model for each biomarker with 5-year prognostic risk score variables (4, 5) dependent, using the appropriate regression method as appropriate: linear regressions for age, FEV_1_% and weight-for-age *z*-score; quasi-Poisson regression for pulmonary exacerbation count in the year prior to enrollment; logistic regressions for pancreatic sufficiency, diabetes and status of infections with MSSA and *Burkholderia cepacia* complex. Because of their clinical importance (3, 21), we evaluated relationships between

### MRSA and *Pseudomonas aeruginosa* status with potential biomarkers using logistic regression

To develop potentially explanatory models for our primary outcome, time to next pulmonary exacerbation, we fitted univariable proportional hazards models (19) for each biomarker, adjusted for confounding variables and excluded mediating variables identified by mediation analysis (11). For each set of models, we accounted for multiple testing using FDR analysis (22) for cutoff values of 0.001, 0.01, 0.05, 0.1 and 0.2 to understand the support for discovered associations for different biomarkers.

### Sensitivity Analyses

We assessed sensitivity of models to anti-inflammatory treatments including inhaled and oral steroids, oral azithromycin and other antibiotics and inhaled antibiotics including aztreonam and tobramycin in any form. We examined ivacaftor or ivacaftor-lumacaftor combination medication effects because they alter airway physiology (3). In each case, we examined the models for independent effects and interactions with potential biomarkers that could indicate mediation of medication effects. We assessed the impact of using Global Lung Initiative equations (23) for estimating FEV_1_% instead of NHANES III equations (15).

### Missingness of Biomarker Measurements

Some sputum samples were small, preventing some biomarker measurements. We preferentially assayed NE, HMGB1 and Calprotectin (12). To evaluate missingness associated bias, we performed multiple imputation by chained equations (24) for completely missing biomarker values using times to next exacerbation, non-missing biomarker values, age, sex, FEV_1_%, weight-for-age *z*-score and number of prior pulmonary exacerbations. We performed three analyses, first, using a dataset with deletion of records with biomarkers that were completely or partially unmeasured, second, using records with deletion of records with completely unmeasured biomarkers but retaining records with partial measurements and third, with all records with imputed values as needed, seeking evidence that missingness was other than completely at random.

## Results

### Patients and Clinical Measurements

We enrolled 114 from Mountain West CF Consortium (MWCFC) patients and collected sputum samples from December 8, 2014 through January 16, 2016. All patients completed follow up to the next pulmonary exacerbation after enrollment or were censored at study end which occurred up to 2.65 years after enrollment.

Characteristics differed between patients who had at least one pulmonary exacerbation or had none within one year post-enrollment. The differences suggested that the 5-year prognostic risk score (4, 5), FEV_1_%, number of exacerbations in the year prior to enrollment and weight-for-age *z*-score are potential clinical confounders, mediators or both for time to next exacerbation (Table 1). No other variable incorporated in the 5-year prognostic risk score or methicillin resistant *Staphylococcus aureus* (MRSA) or *Pseudomonas aeruginosa* significantly differed between patients with and without an exacerbation during one year follow up.

**Table 1.** Enrollment Characteristics and Treatments for Patients by Exacerbations Observed Within One Year.

| Characteristic or Chronic Treatment Received | All Patients<br>n = 114 | Exacerbations Observed in One Year |  | p value |
| --- | --- | --- | --- | --- |
|  |  | None<br>n = 42 | One or More<br>n = 72 |  |
| Years of Age* | 26.3 (12.8 to 68.2) | 27.8 (12.8 to 68.2) | 24.5 (13 to 67.1) | 0.084<sup>§</sup> |
| FEV<sub>1</sub>%* | 72.1 (19.6 to 119) | 80.5 (30.2 to 119) | 66.5 (19.6 to 113) | 0.018<sup>§</sup> |
| Exacerbations in Year Prior to Enrollment* | 1 (0 to 7) | 0 (0 to 5) | 2 (0 to 7) | <0.001<sup>¶</sup> |
| Weight-for Age z-score* | -0.118 (-3.29 to 2.72) | -0.0257 (-1.41 to 2.12) | -0.147 (-3.29 to 2.72) | 0.23<sup>§</sup> |
| Female<sup>†</sup> | 0.46 | 0.43 | 0.49 | 0.69<sup>*</sup> |
| Diabetes<sup>†</sup> | 0.22 | 0.095 | 0.29 | 0.027<sup>*</sup> |
| Pancreatic Sufficiency<sup>†</sup> | 0.079 | 0.12 | 0.056 | 0.29<sup>*</sup> |
| MSSA Infection<sup>†</sup> | 0.45 | 0.48 | 0.43 | 0.78<sup>*</sup> |
| *Burkholderia cepacia* Complex Infection<sup>†</sup> | 0.026 | 0 | 0.042 | 0.30<sup>‡</sup> |
| 5-Year Prognostic Risk Score* | 3.16 (-1.8 to 6.08) | 3.73 (0.676 to 6.08) | 2.76 (-1.8 to 5.59) | <0.001<sup>§</sup> |
| Inhaled Steroids | 0.59 | 0.5 | 0.64 | 0.21<sup>*</sup> |
| Oral Steroids | 0.07 | 0.12 | 0.042 | 0.14<sup>*</sup> |
| Any Steroids | 0.61 | 0.55 | 0.64 | 0.45<sup>*</sup> |
| Azithromycin | 0.54 | 0.48 | 0.58 | 0.36<sup>*</sup> |
| Other Oral Antibiotic | 0.11 | 0.095 | 0.12 | 0.77<sup>‡</sup> |
| Any Oral antibiotics | 0.61 | 0.5 | 0.67 | 0.12<sup>*</sup> |
| Inhaled Tobramycin | 0.33 | 0.31 | 0.35 | 0.84<sup>*</sup> |
| Inhaled Aztreonam | 0.35 | 0.29 | 0.39 | 0.36<sup>*</sup> |
| Any Inhaled Antibiotics | 0.61 | 0.52 | 0.67 | 0.19<sup>*</sup> |
| Any Antibiotics | 0.82 | 0.74 | 0.86 | 0.17<sup>*</sup> |
| Ivacaftor | 0.044 | 0.071 | 0.028 | 0.36<sup>‡</sup> |
| Ivacaftor-Lumacaftor | 0.018 | 0 | 0.028 | 0.53<sup>‡</sup> |
| Ivacaftor or Ivacaftor-lumacaftor | 0.061 | 0.071 | 0.056 | 0.71<sup>‡</sup> |

\*Median (Range)
<sup>†</sup>Decimal Fraction for Status
<sup>§</sup>Linear Regression
<sup>¶</sup>Quasipoisson Regression
<sup>\*</sup>χ-square Test
<sup>‡</sup>Fisher's Exact Test

Kaplan-Meier analysis, logistic regression and proportional hazards modeling found that FEV_1_%, number of prior pulmonary exacerbations, weight-for-age *z*-score and diabetes were associated with time to next exacerbation within a year of enrollment (Supplemental Figure 1, Supplemental Table 3A-B). Proportional hazards modeling confirmed these associations (Supplemental Tables 3B-C) and confirmed these specific clinical markers as potential confounders, mediators or both (10, 11). The remaining prognostic risk score variables, MRSA or *Pseudomonas aeruginosa* infection status were not associated with time to next exacerbation.

### Biomarkers

Biomarker measurements outside the limits of detection were adjusted after log transformation (Supplemental Table 4), to preserve partial information and retain as many patients as possible for analysis. Multiple correlations between biomarkers remained significant after Bonferroni correction (Supplemental Table 5) suggesting that confounding and mediation may exist among the biomarkers themselves.

### Biomarkers, Confounding and Mediation Analyses

We found multiple univariable associations among biomarkers by proportional hazards modeling for time to next pulmonary exacerbation. The results suggest that some potential biomarkers may be causal variables for time to next pulmonary exacerbation (11) (Table 2 and Supplemental Table 6).

**Table 2.** Univariable Biomarker Proportional Hazards Models of Time to Next Exacerbation^*^.

| **Biomarker** | **Estimate** | **SE** | **Hazard Ratio** | ***p*** | **95% CI of the Hazard Ratio** |
| --- | --- | --- | --- | --- | --- |
| MPO | 0.24 | 0.10 | 1.28 | 0.01 | 1.06 to 1.54 |
| NE | 0.18 | 0.08 | 1.20 | 0.02 | 1.03 to 1.39 |
| S100A9 | -0.27 | 0.12 | 0.76 | 0.03 | 0.6 to 0.971 |
| sRAGE | 0.20 | 0.09 | 1.22 | 0.03 | 1.02 to 1.45 |
| ENRAGE | 0.20 | 0.09 | 1.22 | 0.03 | 1.02 to 1.46 |
| IL1β | 0.16 | 0.08 | 1.17 | 0.04 | 1.01 to 1.37 |
| YKL40 | 0.19 | 0.10 | 1.21 | 0.06 | 0.996 to 1.46 |
| IL17A | 0.30 | 0.17 | 1.35 | 0.08 | 0.967 to 1.88 |
| PR3 | 0.14 | 0.09 | 1.15 | 0.11 | 0.967 to 1.37 |
| ICAM1 | 0.24 | 0.15 | 1.27 | 0.12 | 0.938 to 1.71 |
| IL8 | 0.36 | 0.24 | 1.44 | 0.13 | 0.902 to 2.29 |
| Calprotectin | 0.05 | 0.04 | 1.06 | 0.13 | 0.984 to 1.13 |
| MMP9 | 0.13 | 0.08 | 1.13 | 0.13 | 0.962 to 1.34 |

\* See Supplemental Table 6 for results for all biomarkers.

We evaluated confounding and mediation by clinical variables (Supplemental Figure 2) and found associations between different subsets of our markers with each clinical variable except sex (Table 3 and Supplemental Table 7A-E). With a false discovery rate (FDR) <0.001, three biomarkers were significantly associated with FEV_1_%, secretory leukoprotease inhibitor (SLPI), NE and myeloperoxidase (MPO), and for an FDR <0.01, an additional three were significant, soluble receptor for advanced glycation end-products (sRAGE), matrix metallopeptidase 9 (MMP9) and extracellular newly identified receptor for advanced glycation end-products (ENRAGE) (Supplemental Figure 3). S100A8 was likely to be associated with weight-for-age *z-*score (Table 3). Altogether, these findings suggest a potential for confounding or mediation by these clinical markers (11).

**Table 3.**
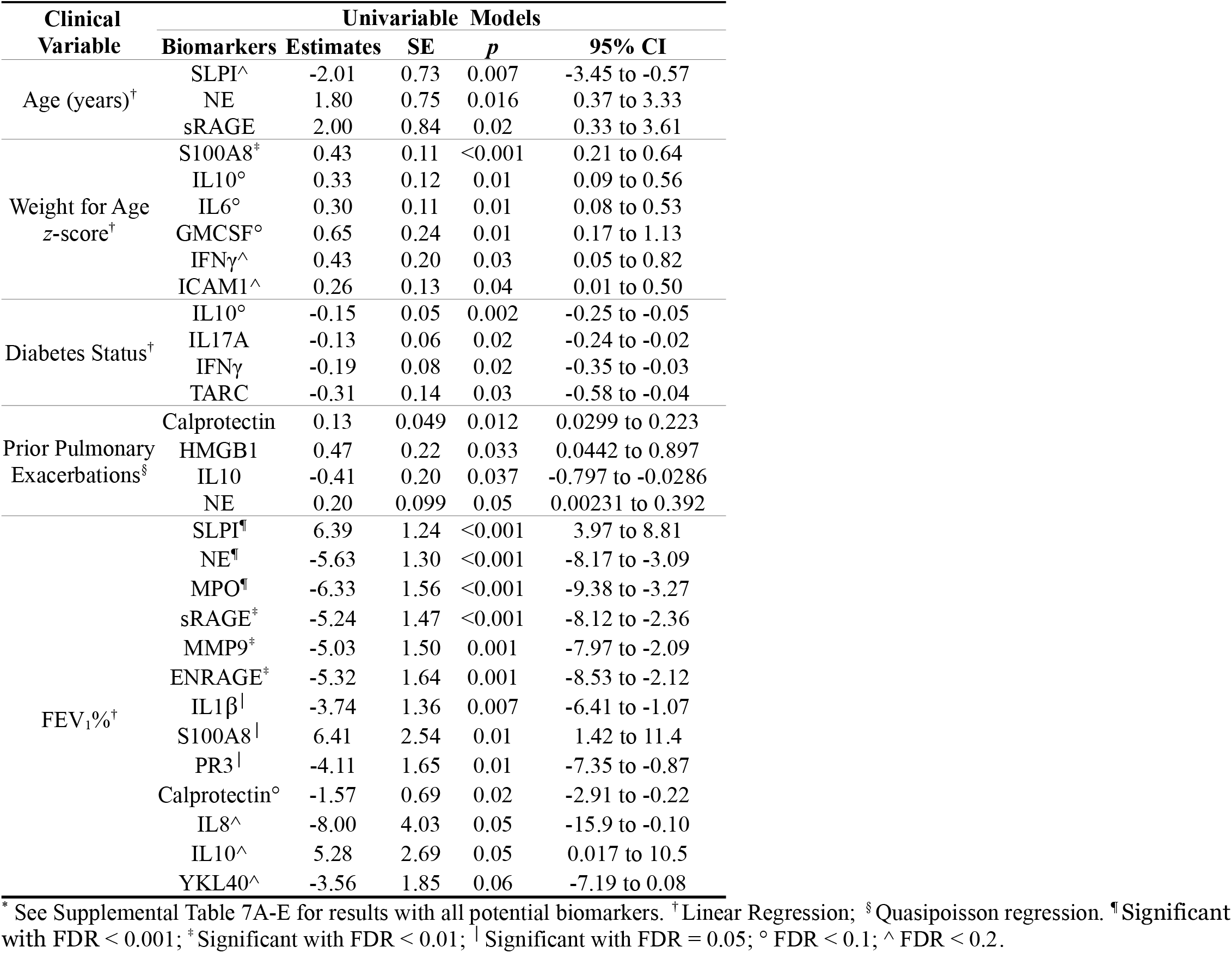
Unadjusted Associations between Biomarkers and Clinical Characteristics.^*^

| Clinical Variable | Univariable Models |  |  |  |  |
| --- | --- | --- | --- | --- | --- |
|  | Biomarkers | Estimates | SE | p | 95% CI |
| Age (years)<sup>†</sup> | SLPI<sup>^</sup> | -2.01 | 0.73 | 0.007 | -3.45 to -0.57 |
|  | NE | 1.80 | 0.75 | 0.016 | 0.37 to 3.33 |
|  | sRAGE | 2.00 | 0.84 | 0.02 | 0.33 to 3.61 |
| Weight for Age z-score<sup>†</sup> | S100A8<sup>‡</sup> | 0.43 | 0.11 | <0.001 | 0.21 to 0.64 |
|  | IL10<sup>°</sup> | 0.33 | 0.12 | 0.01 | 0.09 to 0.56 |
|  | IL6<sup>°</sup> | 0.30 | 0.11 | 0.01 | 0.08 to 0.53 |
|  | GMCSF<sup>°</sup> | 0.65 | 0.24 | 0.01 | 0.17 to 1.13 |
| | IFNγ<sup>^</sup> | 0.43 | 0.20 | 0.03 | 0.05 to 0.82 |
|  | ICAM1<sup>^</sup> | 0.26 | 0.13 | 0.04 | 0.01 to 0.50 |
| Diabetes Status<sup>†</sup> | IL10<sup>°</sup> | -0.15 | 0.05 | 0.002 | -0.25 to -0.05 |
|  | IL17A | -0.13 | 0.06 | 0.02 | -0.24 to -0.02 |
| | IFNγ | -0.19 | 0.08 | 0.02 | -0.35 to -0.03 |
|  | TARC | -0.31 | 0.14 | 0.03 | -0.58 to -0.04 |
| Prior Pulmonary Exacerbations<sup>§</sup> | Calprotectin | 0.13 | 0.049 | 0.012 | 0.0299 to 0.223 |
|  | HMGB1 | 0.47 | 0.22 | 0.033 | 0.0442 to 0.897 |
|  | IL10 | -0.41 | 0.20 | 0.037 | -0.797 to -0.0286 |
|  | NE | 0.20 | 0.099 | 0.05 | 0.00231 to 0.392 |
| FEV<sub>1</sub>%<sup>†</sup> | SLPI<sup>¶</sup> | 6.39 | 1.24 | <0.001 | 3.97 to 8.81 |
|  | NE<sup>¶</sup> | -5.63 | 1.30 | <0.001 | -8.17 to -3.09 |
|  | MPO<sup>¶</sup> | -6.33 | 1.56 | <0.001 | -9.38 to -3.27 |
|  | sRAGE<sup>‡</sup> | -5.24 | 1.47 | <0.001 | -8.12 to -2.36 |
|  | MMP9<sup>‡</sup> | -5.03 | 1.50 | 0.001 | -7.97 to -2.09 |
|  | ENRAGE<sup>‡</sup> | -5.32 | 1.64 | 0.001 | -8.53 to -2.12 |
| | IL1β<sup>‡</sup> | -3.74 | 1.36 | 0.007 | -6.41 to -1.07 |
|  | S100A8<sup>‡</sup> | 6.41 | 2.54 | 0.01 | 1.42 to 11.4 |
|  | PR3<sup>‡</sup> | -4.11 | 1.65 | 0.01 | -7.35 to -0.87 |
|  | Calprotectin<sup>°</sup> | -1.57 | 0.69 | 0.02 | -2.91 to -0.22 |
|  | IL8<sup>^</sup> | -8.00 | 4.03 | 0.05 | -15.9 to -0.10 |
|  | IL10<sup>^</sup> | 5.28 | 2.69 | 0.05 | 0.017 to 10.5 |
|  | YKL40<sup>^</sup> | -3.56 | 1.85 | 0.06 | -7.19 to 0.08 |

\* See Supplemental Table 7A-E for results with all potential biomarkers. <sup>†</sup>Linear Regression; <sup>§</sup>Quasipoisson regression. <sup>¶</sup>Significant with FDR < 0.001; <sup>‡</sup>Significant with FDR < 0.01; <sup>‡</sup>Significant with FDR = 0.05; <sup>°</sup> FDR < 0.1; <sup>^</sup> FDR < 0.2.

By definition, age, sex, and prior pulmonary exacerbations, cannot mediate associations between biomarkers and time to next exacerbation. Thus, we treated them as confounders in further modeling with biochemical markers of inflammation.

We evaluated weight-for-age *z*-score, diabetes and FEV_1_% as potential confounders, mediators or both. In proportional hazards models of weight-for-age *z*-score and diabetes for time to next exacerbation, the hazard ratios and *p*-values for the biomarkers used as adjustment variables (Table 4 and Supplemental Tables 8A-C) were similar to those found in earlier models of the biomarkers alone for time to next exacerbation (Table 2 and Supplemental Table 6). Weight-for-age *z*-score and diabetes are unlikely to be confounders or mediators of inflammation.

**Table 4.** Biomarkers as Univariable Adjustments to Clinical Variables in Proportional Hazards Models of Time to Next Exacerbation.

| Clinical Variable* | Adjustment Univariables |  |  |  |  |  |
| --- | --- | --- | --- | --- | --- | --- |
|  | Biomarkers | Estimates | SE | Hazard Ratio | p | 95% CI of the Hazard Ratio |
| Weight for Age z-score | MPO^ | 0.29 | 0.09 | 1.33 | 0.002 | 1.11 to 1.6 |
|  | NE | 0.17 | 0.07 | 1.18 | 0.03 | 1.02 to 1.37 |
|  | S100A9 | -0.27 | 0.12 | 0.76 | 0.03 | 0.597 to 0.971 |
|  | sRAGE | 0.19 | 0.09 | 1.21 | 0.04 | 1.01 to 1.44 |
|  | ENRAGE^ | 0.25 | 0.09 | 1.29 | 0.01 | 1.07 to 1.55 |
|  | IL1β | 0.17 | 0.08 | 1.18 | 0.03 | 1.02 to 1.37 |
|  | YKL40 | 0.19 | 0.09 | 1.20 | 0.05 | 1 to 1.45 |
|  | IL17A | 0.43 | 0.18 | 1.54 | 0.02 | 1.08 to 2.2 |
|  | PR3 | 0.12 | 0.09 | 1.13 | 0.16 | 0.952 to 1.33 |
|  | ICAM1 | 0.32 | 0.16 | 1.38 | 0.04 | 1.02 to 1.87 |
|  | IL8 | 0.42 | 0.24 | 1.52 | 0.08 | 0.956 to 2.41 |
|  | Calprotectin | 0.06 | 0.04 | 1.06 | 0.10 | 0.989 to 1.14 |
|  | MMP9 | 0.16 | 0.09 | 1.17 | 0.07 | 0.989 to 1.39 |
| Diabetes Status | MPO | 0.22 | 0.10 | 1.24 | 0.03 | 1.03 to 1.5 |
|  | NE | 0.17 | 0.08 | 1.19 | 0.03 | 1.02 to 1.38 |
|  | S100A9 | -0.25 | 0.13 | 0.78 | 0.05 | 0.607 to 0.995 |
|  | sRAGE | 0.19 | 0.09 | 1.21 | 0.03 | 1.02 to 1.45 |
|  | ENRAGE | 0.18 | 0.09 | 1.19 | 0.06 | 0.993 to 1.43 |
|  | IL1β | 0.15 | 0.08 | 1.17 | 0.05 | 0.999 to 1.36 |
|  | YKL40 | 0.17 | 0.10 | 1.19 | 0.09 | 0.975 to 1.44 |
|  | IL17A | 0.30 | 0.16 | 1.34 | 0.07 | 0.973 to 1.86 |
|  | PR3 | 0.12 | 0.09 | 1.13 | 0.17 | 0.946 to 1.35 |
|  | ICAM1 | 0.24 | 0.16 | 1.27 | 0.12 | 0.939 to 1.72 |
|  | IL8 | 0.28 | 0.23 | 1.32 | 0.23 | 0.839 to 2.09 |
|  | Calprotectin | 0.06 | 0.04 | 1.06 | 0.11 | 0.986 to 1.14 |
|  | MMP9 | 0.11 | 0.08 | 1.12 | 0.19 | 0.946 to 1.32 |
| FEV<sub>1</sub>% | MPO | 0.19 | 0.10 | 1.21 | 0.06 | 0.99 to 1.48 |
|  | NE | 0.14 | 0.08 | 1.15 | 0.09 | 0.98 to 1.35 |
|  | S100A9 | -0.24 | 0.12 | 0.79 | 0.05 | 0.619 to 1 |
|  | sRAGE | 0.14 | 0.10 | 1.15 | 0.14 | 0.954 to 1.39 |
|  | ENRAGE | 0.15 | 0.10 | 1.16 | 0.12 | 0.961 to 1.41 |
|  | IL1β | 0.13 | 0.08 | 1.14 | 0.11 | 0.973 to 1.33 |
|  | YKL40 | 0.15 | 0.10 | 1.16 | 0.14 | 0.951 to 1.41 |
|  | IL17A | 0.24 | 0.17 | 1.28 | 0.16 | 0.908 to 1.79 |
|  | PR3 | 0.11 | 0.09 | 1.12 | 0.22 | 0.935 to 1.35 |
|  | ICAM1 | 0.18 | 0.15 | 1.20 | 0.23 | 0.888 to 1.63 |
|  | IL8 | 0.29 | 0.24 | 1.33 | 0.23 | 0.83 to 2.14 |
|  | Calprotectin | 0.04 | 0.04 | 1.04 | 0.34 | 0.963 to 1.11 |
|  | MMP9 | 0.08 | 0.09 | 1.08 | 0.37 | 0.909 to 1.29 |

\* See Supplemental Table 8A-C for results of all biomarkers. † Significant with FDR = 0.001; \* Significant with FDR = 0.01; ‡ Significant with FDR = 0.05; ° FDR < 0.1; ^ FDR < 0.2.

Estimated biomarker hazard ratios were weaker and uniformly non-significant when used as adjustments in proportional hazards models of FEV_1_% for time to next exacerbation (Table 4 and Supplemental Tables 8A-C). FEV_1_% partially mediates the association between inflammation and time to next exacerbation and should be excluded from further models (11).

### Biomarkers and Pulmonary Exacerbations

Multivariable proportional hazards models of each potential biomarker including age, sex and prior pulmonary exacerbations as confounders for time to next pulmonary exacerbation (10) found 14 inflammatory markers with hazard ratios for time to next exacerbation with *p* < 0.2 (Table 5 and Supplemental Table 9).

**Table 5.** Biomarkers in Adjusted^*^ Proportional Hazards Models of Time to Next Exacerbation.

| Biomarkers | Estimates | SE | Hazard Ratio | *p* | 95% CI of the Hazard Ratio |
| --- | --- | --- | --- | --- | --- |
| ENRAGE<sup>°</sup> | 0.30 | 0.10 | 1.35 | 0.004 | 1.1 to 1.65 |
| MPO<sup>°</sup> | 0.26 | 0.099 | 1.30 | 0.008 | 1.07 to 1.57 |
| sRAGE<sup>^</sup> | 0.20 | 0.088 | 1.23 | 0.021 | 1.03 to 1.46 |
| ICAM1<sup>^</sup> | 0.38 | 0.16 | 1.46 | 0.021 | 1.06 to 2.02 |
| NE<sup>^</sup> | 0.18 | 0.077 | 1.19 | 0.022 | 1.03 to 1.39 |
| YKL40<sup>^</sup> | 0.23 | 0.10 | 1.26 | 0.024 | 1.03 to 1.53 |
| TARC<sup>^</sup> | 1.00 | 0.46 | 2.72 | 0.031 | 1.10 to 6.74 |
| MMP9<sup>^</sup> | 0.18 | 0.09 | 1.20 | 0.039 | 1.01 to 1.43 |
| IL1B<sup>^</sup> | 0.16 | 0.08 | 1.17 | 0.049 | 1.00 to 1.37 |
| IL5<sup>^</sup> | 0.58 | 0.31 | 1.79 | 0.062 | 0.97 to 3.29 |
| IL17A | 0.28 | 0.17 | 1.32 | 0.098 | 0.95 to 1.83 |
| S100A9 | -0.20 | 0.12 | 0.823 | 0.12 | 0.645 to 1.05 |
| IFNγ | 0.37 | 0.26 | 1.44 | 0.16 | 0.861 to 2.41 |
| PR3 | 0.12 | 0.094 | 1.13 | 0.19 | 0.94 to 1.36 |

\* Adjusted by Age, Sex and Number of Prior Pulmonary Exacerbations. See Supplemental Table 9 for results for all biomarkers.
<sup>¶</sup> Significant with FDR = 0.001; <sup>\*</sup> FDR = 0.01; <sup>†</sup> FDR = 0.05; <sup>°</sup> FDR < 0.1; <sup>^</sup> FDR < 0.2.

Among these, setting false discovery rate (FDR) < 0.2 sets an 80% likelihood suggesting eight of the first ten have true associations with time to next exacerbation (Supplemental Figure 4). Setting the FDR < 0.01 shows ENRAGE and MPO each have > 99% chance of being true findings (Table 5).

### Sensitivity Analyses

Adding inflammation modifying treatments as confounders to the proportional hazards models adjusted by age, sex and number of prior exacerbations did not substantially modify coefficients for any biomarker. Use of corticosteroids, inhaled, oral or both, chronic azithromycin, inhaled aztreonam, tobramycin, each alone or alternating every other month had no significant independent associations with time to next exacerbation nor interactions with the biomarker variables in each model. Coefficients (Table 5) remained stable throughout sensitivity testing suggesting that results are not due to anti-inflammatory treatments.

Seven patients used ivacaftor or ivacaftor-lumacaftor, but their use had no independent effect or interaction, and individual biomarker variable effects were stable throughout testing. Use of Global Lung Initiative equations (23) to calculate FEV_1_% instead of NHANES III led to similar results and identical interpretations and conclusions.

### Biomarker Measurement Missingness

We repeated models to understand the impact of partially missing information due to biomarker measurements outside detection limits and completely missing data due to insufficient samples. Analyses excluding patients with partially missing data produced similar results for models derived from using complete data sets. Analyses with missing data imputed using multiple imputation with chained equations produced similar model results from using complete data sets (Supplemental Tables 6-9). We found no evidence of bias for any relationship examined.

## Discussion

We measured 24 sputum biomarkers from 114 randomly chosen, clinically stable adolescents and adults with CF to validate prior observations for calprotectin, NE and HMGB1 (6–8) and to explore causal inference, confounding (10) and mediation (11) between an expanded list of inflammatory biomarkers and clinical outcomes. After evaluation for confounders and mediators of biomarker effects, we found potential associations with ten different molecules: ENRAGE, MPO, sRAGE, ICAM1, NE, chitinase-3-like 1 protein (YKL40), thymus and activation regulated chemokine (TARC), MMP9, IL1β and IL5 (Table 5). At least eight are associated with time to next pulmonary exacerbation (Supplemental Figure 4). FEV_1_% mediates biomarker effects and was excluded from every model, while age, sex, number of prior pulmonary exacerbations were confounders and were included (10, 11). Results were insensitive to anti-inflammatory treatments or ivacaftor, although the latter was infrequently used.

Severe CF lung disease continues to steadily decrease as treatments with CFTR modulators increase (25). FEV_1_% remains the single most common measure of disease severity, but it strongly mediates and obscures biomarker effects (Table 4). FEV_1_% poorly distinguishes between rapid or slow worsening of lung disease (4). As CF disease severity lessens (26), FEV_1_% will become less useful, and biomarkers measuring inflammation with appropriate adjustments for confounding will be increasingly useful for understanding clinical status (Figure 1) and providing attractive intervention targets.

**Figure 1.**
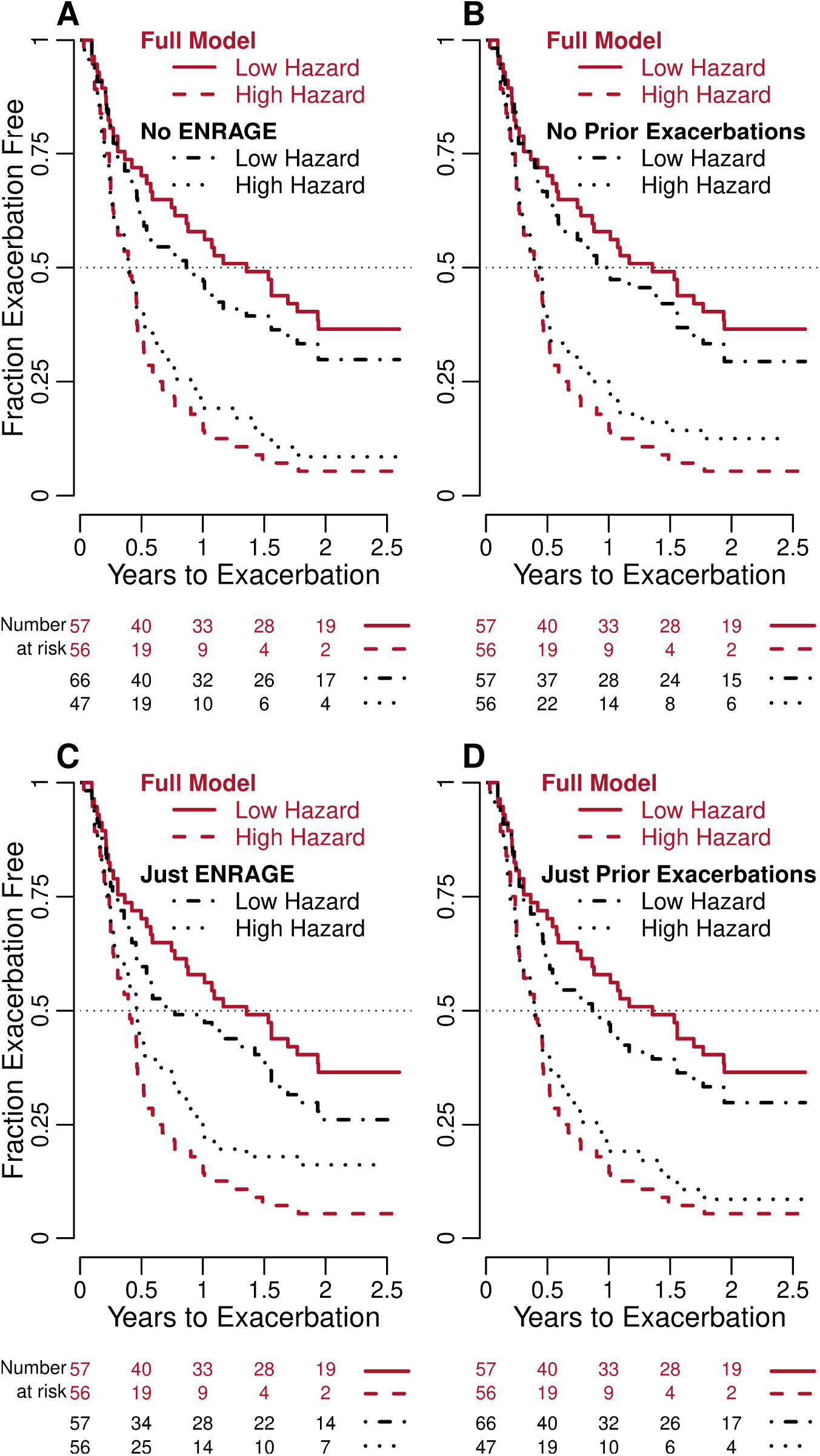
Kaplan Meier Plots Based on Proportional Hazard Ratio Estimates for Individual Patients. We categorized patients by hazard ratios higher or lower than the median estimated using a **full model** including log transformed values of ENRAGE adjusted by age, sex and number of prior pulmonary exacerbations as confounders. We drew Kaplan-Meier plots and compared with plots based on hazard ratios estimated using model **A. excluding ENRAGE data, B. excluding prior pulmonary exacerbations, C. including only ENRAGE data**, and **D. including only prior pulmonary exacerbations**. The numbers at risk for **A** and **D** differ from other plots because median hazard ratios including exacerbation count data did not divide patients into equal groups.

Biomarkers associated with time to next exacerbation fall into several categories (Figure 2). ENRAGE and sRAGE are constituents of RAGE-axis related inflammation. ENRAGE is a high avidity RAGE ligand that indicates neutrophil activity (27). It is elevated in CF airways in infancy (28) and is associated with low FEV_1_ and CF-related diabetes (29). ENRAGE increases during pulmonary exacerbation and falls with antibiotic treatment (30). Beyond CF, higher ENRAGE is associated with cigarette smoking-induced injury (31), higher coronary artery disease risk (32) and higher chronic kidney disease mortality (33). ENRAGE increases in bronchoalveolar lavage in the acute respiratory distress syndrome (ARDS) and decreases with recovery (34). In SARS-CoV-2-associated ARDS, it rises with increasing severity and falls with recovery (35).

**Figure 2.**
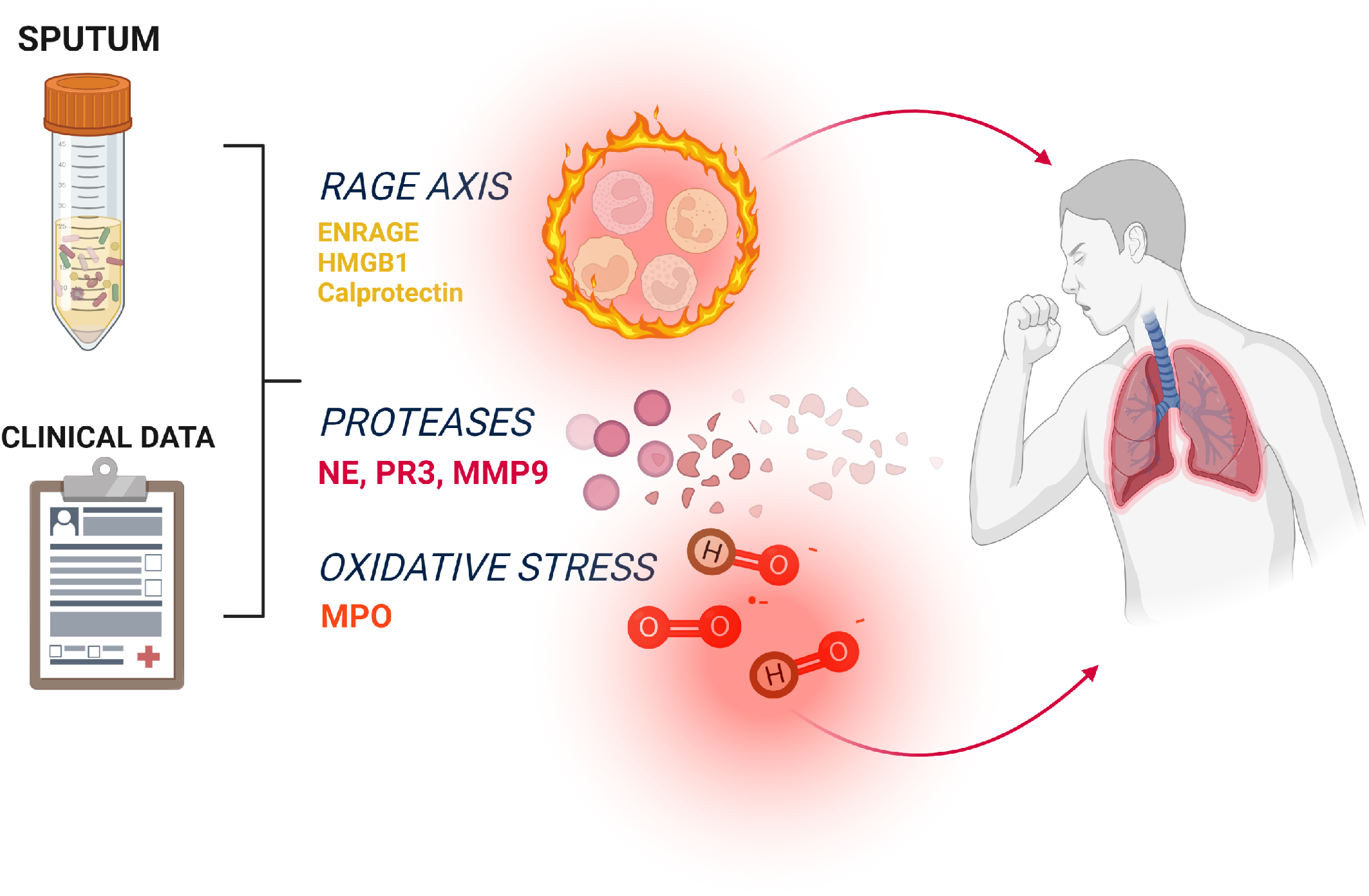
Schematic Model of Inflammation in the CF Airway. Using sputum and clinical data, we measured potential inflammatory biomarkers to investigate pulmonary exacerbations of CF. We found that RAGE Axis related inflammation and neutrophil related protease-anti-protease imbalance and oxidative stress are the most likely pathophysiologic processes involved.

ENRAGE most strongly correlated with MMP9 and MPO (Supplemental Table 5) suggesting associations between RAGE axis inflammation and both protease and oxidative stress mediated injuries. SRAGE is the soluble form of the membrane bound RAGE receptor. Strong correlations with NE (0.80), MMP9 (0.65) and IL1β (0.69) may indicate associations between RAGE axis inflammation and neutrophil-derived protease mediated injuries. MPO, ICAM1, NE, YKL40, TARC and MMP9 reflect the important roles of neutrophils, neutrophil-associated proteases (MMP9) (7, 36) and reactive oxygen species (MPO) (37) in CF airway inflammation (Figure 2).

SLPI, NE and MPO (Supplemental Figure 3) were associated with FEV_1_% with a FDR set at 0.001. For FDR set at 0.01, we found another three biomarkers, sRAGE, MMP9 and ENRAGE. Anti-protease SLPI opposes NE, MMP9 and proteinase 3 (PR3) activities (7, 37, 38) and was associated with higher FEV_1_% (Table 3). These findings suggest that RAGE-axis, protease-anti-protease imbalance and reactive oxygen species sources of inflammation might cause lower FEV_1_% (Supplemental Figure 5).

Contrary to pre-study expectations (6–8), Calprotectin, NE and HMGB1 were not associated with time to next pulmonary exacerbation. However, the study redemonstrated associations between these molecules and prior pulmonary exacerbations (Table 3 and Supplemental Table 7D), and they are markers of neutrophil activity and part of the RAGE-axis of inflammation. These partial validation results reinforce that this study differs from prior studies in at least two substantial ways. First, we recruited randomly selected clinically stable rather than mixed stable and unstable serial patients. Second, our analyses evaluated confounding and mediation by clinical covariables (10, 11).

Our study has limitations. Because we are pursuing explanatory airway inflammation models (13), no methods of model selection are sufficient (10) to confidently produce a multivariable panel of inflammatory biomarkers. However, our current observational work will be useful prior to launching new observational and other types of investigations to discover a multivariable biomarker model. Although many study patients took every prior effective treatment, due to study timing, only 6% received CFTR modulators. Our study does not confirm whether CFTR modulators reduce airway inflammation. Further studies are needed, however, modulators are associated with reduced sputum production, so alternative sampling methods may be required.

Finally, our explanatory models infer but cannot confirm causal relationships. RAGE axis inflammation, protease mediated injury and oxidative stress are strongly associated with future exacerbation. FEV_1_%, the single most important predictor of survival in CF, mediates the association of inflammatory markers with time to next pulmonary exacerbation. Further studies in cell culture or animal models or human trials of anti-proteases, anti-oxidants, RAGE inhibiting agents or a combination are needed to provide the information to improve causal inference and potentially treat the CF airway inflammation. Because of the importance and commonality of these inflammatory processes in other lung diseases such as ARDS, progress in CF with novel anti-inflammatory treatments may have broad application.

## Data Availability

All data produced in the present study will be available upon reasonable request to the authors after peer-reviewed publication.

## Author Contributions

TGL is the manuscript guarantor and takes responsibility for the integrity of the work from inception to publication. TGL drafted the initial manuscript; all authors contributed interpretive insights, edits and corrections. MH created the graphical abstract with assistance from TGL and assisted TGL in turn to diagram interactions between biomarkers and lung disease. DRC initiated and oversaw development of the statistical plan for randomized enrollment by TGL, FRA, RK with additional inputs from JLJ, KRP and SDS. TGL, FRA, DRC, CK, RK and RDS performed the statistical analysis. JLJ oversaw IRB submissions throughout the MWCFC with help at each site from NA, DD, JAF, BG, CMK, OM, KAP, KRP, AJR, JBV and RY. TGL, NA, PSB, BAC, CLD, DD, JAF, BG, JLJ, TH, JRH, CMK, YL, JL, NL, OM, CN, KAP, KRP, ALQ, PR, AJR, SDS, JLT-C, JBV, RY, JPC, JSE, KNO developed and performed patient enrollment processes, sample collections and clinical annotations. JLJ oversaw the work preceding the study planning meeting with critical assistance from KAP, JAF and JBV. JLJ oversaw and coordinated logistics and regulatory documentation for the entire study.

TGL, JAF, JLJ, YL, KAP and JBV developed the sputum processing protocol. JAF, JLJ, KAP, YL and JBV provided central and on-site laboratory training. TGL and JLJ performed site initiation visits. JLJ and KAP centrally monitored data collection. JAF, KAP and JBV performed interim and end-of-study site visits to verify data integrity and regulatory compliance. KAP developed and implemented REDCap electronic data capture with input from TGL, JAF, JLJ and JBV. KAP oversaw data entry, query generation and resolution and generation of raw data reports. JBV with assistance from JAF, JLJ and YL developed protocols and oversaw specimen shipping by NA, DD, BG, CMK, OM, KRP, AJR and RY. JAF, JLJ, JBV, KAP and YL received, handled and successfully managed sputum specimens centrally despite shipment damage, floods, power outages, freezer and complete building services failures. YL performed ELISA studies in Utah. SS oversaw NE activity determinations in Colorado. TGL obtained funding with assistance from FRA, JLJ and SDS and additional help from PSB, BAC, CLD, RK, JL, CN and DRC. JPC, JSE and KNO were the Study Advisory Committee and contributed to multiple study aspects. All authors had access to complete raw and processed datasets from this study.

The results, interpretations and opinions expressed in this work are those of the authors and do not necessarily represent the views of their institutions, the NIH, NHLBI, NCATS, NSF, the Veteran’s Administration, the Margolis Foundation, the Claudia Ruth Goodrich Stevens Family, the US CF Foundation, the European CF Society or the Medical Research Council in the UK or any of the sponsors of clinical trials that provided other support for individual authors or the governments of the US or the UK.

## Acknowledgments

We thank the study participants and their families. We thank Mike Mitchell in the Pulmonary Laboratory at the University of Utah for technical assistance.

