## Supplemental Table 1 for "Associations of Sputum Biomarkers with Clinical Outcomes in People with Cystic Fibrosis"

**Supplemental Table 1. Investigational Review Board Approvals**

| CF Care Center | Reviewing Entity | Protocol Designation |
|---|---|---|
| University of Utah Adult Care Center and the Intermountain CF Center at Primary Children’s Medical Center | University of Utah IRB | IRB_00011571 |
| St. Luke’s Health System | St. Luke’s Health System IRB | 13-0547 |
| Las Vegas CF Center | The Western IRB | STUDY NUM 1146159<br>WIRB PRO NUM: 20140721<br>INVEST NUM: 112264<br>WO NUM: 1-837046-1 |
| National Jewish Health | National Jewish Health IRB | Protocol Number 20130530<br>Study Number: HS-2797 |
| Children’s Hospital Colorado | Colorado Multiple IRB | COMIRB Protocol 13-3172 |
| Phoenix Children’s Hospital | Phoenix Children’s Hospital IRB | PCH IRB #13-087 |
| University of Arizona, Tucson | University of Arizona IRB | University of Arizona IRB |
| University of New Mexico | Human Research Review Committee in the Human Research Protections Office at the University of New Mexico | Study ID 14-085 |
