## Supplemental Table 2 for "Associations of Sputum Biomarkers with Clinical Outcomes in People with Cystic Fibrosis"

**Supplemental Table 2. Reagents for ELISA Testing***

**Standard Curves**

| Recombinant Human Protein | Source and Catalog Number | Curve Range |
|---|---|---|
| C-Reactive Protein (CRP) | Abcam ab167710 | 15.625 to 1000 pg/ml |
| Calprotectin or S100A8/A9 Heterodimer | Biolegend 753404 | 3.9 to 250 ng/ml |
| High Mobility Group Box-1 (HMGB1) | Sigma-Aldrich H4652 | 3.9 to 250 ng/ml |

**Capture Antibody**

| Source and Target Species of IgG | Source and Catalog Number | Concentration Used |
|---|---|---|
| Rabbit polyclonal anti-human CRP | Abcam ab31156 | 2.5 µg/ml |
| Mouse monoclonal anti-human Calprotectin | LS-Bio LS-C96223 | 1.5 µg/ml |
| Rabbit polyclonal anti-human HMGB1 | Upstate Biotechnology 07-584 | 1.5 µg/ml |

**Detection Antibody**

| Species | Source and Catalog Number | Concentration or Dilution Used |
|---|---|---|
| Mouse monoclonal IgG anti-human CRP | Abcam ab136176 | 1.5 µg/ml |
| Rabbit polyclonal anti-human Calprotectin | LS-Bio LS122793 | 1:1500 |
| Mouse monoclonal anti-human HMGB1 | R&D Systems MAB1690 | 0.75 µg/ml FC |

**Enzyme Linked Antibody**

| Species | Source and Catalog Number | Dilution Used |
|---|---|---|
| Goat Anti-Mouse Polyclonal IgG-FC | Millipore AP127P | 1:2000 |
| Goat Anti-Rabbit IgG-FC | Santa Cruz Biotechnology SC-2004 | 1:2000 |
| Goat Anti-Mouse Polyclonal IgG-FC | Millipore AP127P | 1:2000 |

***** 96-well plates (Costar, Corning Inc, Costar, NY, USA) were incubated overnight with capture antibody diluted according to manufacturer recommendations or 1% bovine serum albumin (BSA, MilliporeSigma, Burlington, MA, USA) or 10% newborn calf serum in phosphate buffered saline (NCS-PBS, MilliporeSigma). Plates were washed 4 times with PBS prior to addition of 1st antibodies then washed 6 times prior to addition of horse radish peroxidase-conjugated 2nd antibodies. Standards were diluted 1:2 for standard curves using manufacturer's recommendations or 1% BSA or 10% NCS-PBS (MilliporeSigma). All assays were incubated with 3,3′,5,5′-Tetramethylbenzidine (TMB) substrate solution (Thermo Scientific, Waltham, MA, USA). Reactions were stopped using 0.18 M $H_2SO_4$ after 15-30 minutes and read in an ELISA plate reader at $OD_{450}$. Standard curves were constructed using linear regression of log-transformed mean fluorescence intensities and log-transformed known protein concentrations.
