## Supplemental Table 3 for "Associations of Sputum Biomarkers with Clinical Outcomes in People with Cystic Fibrosis"

**Supplemental Table 3A. Associations with Exacerbation Within 1 Year of Enrollment by Logistic Regression.**[*]

| Clinical Univariable | Estimate | Standard Error (SE) | *p* | 95% CI |
|---|---|---|---|---|
| Age (years) | -0.028 | 0.016 | 0.09 | -0.059 to 0.0043 |
| Sex (Male=0, Female=1) | 0.23 | 0.39 | 0.55 | -0.53 to 1.00 |
| $FEV_1$% | -0.022 | 0.0095 | 0.02 | -0.041 to -0.0034 |
| Number of Prior Pulmonary Exacerbations | 0.64 | 0.17 | <0.001 | 0.30 to 0.98 |
| Diabetes Status | 1.36 | 0.59 | 0.02 | 0.22 to 2.50 |
| *Burkholderia cepacia* Complex Infection Status | 16 | 1400 | 0.99 | -2700 to 2700 |
| Weight-for-age *z*-score | -0.24 | 0.20 | 0.23 | -0.64 to 0.16 |
| Methicillin Sensitive *Staphylococcus aureus* Infection Status | -0.18 | 0.39 | 0.64 | -0.95 to 0.58 |
| Pancreatic Sufficiency | -0.83 | 0.70 | 0.24 | -2.20 to 0.54 |
| Methicillin Resistant *S aureus* Infection Status | 0.75 | 0.55 | 0.18 | -0.34 to 1.80 |
| *Pseudomonas aeruginosa* Infection Status | 0.76 | 0.40 | 0.06 | -0.026 to 1.50 |

[*] Univariable Logistic Regression. There was no missingness to consider among these clinical variables.

**Supplemental Table 3B. Associations with Exacerbation Within 1 Year of Enrollment by Proportional Hazards Modeling.**[*]

| Clinical Univariable | Estimate | SE | Hazard Ratio | *p* | 95% CI of the Hazard Ratio |
|---|---|---|---|---|---|
| Age (years) | -0.023 | 0.012 | 0.98 | 0.044 | 0.96 to 1.00 |
| Sex (Male=0, Female=1) | 0.16 | 0.24 | 1.20 | 0.49 | 0.74 to 1.90 |
| $FEV_1$% | -0.015 | 0.006 | 0.99 | 0.009 | 0.97 to 1.00 |
| Number of Prior Pulmonary Exacerbations | 0.32 | 0.071 | 1.40 | <0.001 | 1.20 to 1.60 |
| Diabetes Status | 0.61 | 0.26 | 1.80 | 0.02 | 1.10 to 3.10 |
| *Burkholderia cepacia* Complex Infection Status | 0.50 | 0.59 | 1.70 | 0.39 | 0.52 to 5.30 |
| Weight-for-age *z*-score | -0.30 | 0.14 | 0.74 | 0.034 | 0.57 to 0.98 |
| Methicillin Sensitive *Staphylococcus aureus* Infection Status | -0.14 | 0.24 | 0.87 | 0.56 | 0.55 to 1.40 |
| Pancreatic Sufficiency | -0.56 | 0.51 | 0.57 | 0.27 | 0.21 to 1.60 |
| Methicillin Resistant *S aureus* Infection Status | 0.39 | 0.28 | 1.50 | 0.17 | 0.85 to 2.60 |
| *Pseudomonas aeruginosa* Infection Status | 0.43 | 0.25 | 1.50 | 0.092 | 0.93 to 2.50 |

[*] Univariable Proportional Hazards Models. There was no missingness to consider among these clinical variables.

**Supplemental Table 3C. Associations with Exacerbation Within Entire Study by Proportional Hazards Modeling.**[*]

| Clinical Univariable | Estimate | SE | Hazard Ratio | *p* | 95% CI of the Hazard Ratio |
|---|---|---|---|---|---|
| Age (years) | -0.024 | 0.01 | 0.98 | 0.022 | 0.96 to 1.00 |
| Sex (Male=0, Female=1) | 0.22 | 0.21 | 1.20 | 0.29 | 0.82 to 1.90 |
| $FEV_1$% | -0.012 | 0.005 | 0.99 | 0.013 | 0.98 to 1.00 |
| Number of Prior Pulmonary Exacerbations | 0.28 | 0.064 | 1.30 | <0.001 | 1.20 to 1.50 |
| Diabetes Status | 0.66 | 0.24 | 1.90 | 0.0062 | 1.20 to 3.10 |
| *Burkholderia cepacia* Complex Infection Status | 0.50 | 0.59 | 1.70 | 0.39 | 0.52 to 5.30 |
| Weight-for-age *z*-score | -0.31 | 0.13 | 0.74 | 0.016 | 0.57 to 0.94 |
| Methicillin Sensitive *Staphylococcus aureus* Infection Status | -0.04 | 0.21 | 0.96 | 0.83 | 0.63 to 1.40 |
| Pancreatic Sufficiency | -0.67 | 0.46 | 0.51 | 0.15 | 0.21 to 1.30 |
| Methicillin Resistant *S aureus* Infection Status | 0.31 | 0.26 | 1.40 | 0.24 | 0.81 to 2.30 |
| *Pseudomonas aeruginosa* Infection Status | 0.22 | 0.22 | 1.20 | 0.32 | 0.81 to 1.9 |

[*] Univariable Proportional Hazards Models. There was no missingness to consider among these clinical variables.
