## Supplemental Table 4 for "Associations of Sputum Biomarkers with Clinical Outcomes in People with Cystic Fibrosis"

**Supplemental Table 4. Log Transformed Biomarker Measurements***

| Biomarker† | Patients | Number of Values | | | Original Units | Raw Data with Missing Values Omitted | | | Adjusted for Immunoassay Limits of Detection | | | Adjusted Data with Missing Values Imputed | | |
|---|---|---|---|---|---|---|---|---|---|---|---|---|---|---|
| | | Missing | Below Limits of Detection | Above Limits of Detection | | Mean | SD | CV | Mean | SD | CV | Mean | SD | CV |
| Calprotectin | 114 | 0 | 0 | 0 | ng/ml | 10.9 | 5.48 | 0.50 | 11.6 | 2.96 | 0.25 | 11.6 | 2.96 | 0.25 |
| CRP | 114 | 0 | 0 | 0 | ng/ml | 10.1 | 3.28 | 0.32 | 12.5 | 0.60 | 0.05 | 12.5 | 0.60 | 0.05 |
| ENRAGE | 113 | 1 | 0 | 0 | pg/ml | 14.5 | 1.28 | 0.088 | 14.5 | 1.21 | 0.08 | 14.5 | 1.21 | 0.08 |
| GMCSF | 112 | 2 | 0 | 0 | pg/ml | 2.78 | 0.37 | 0.13 | 2.78 | 0.37 | 0.13 | 2.78 | 0.37 | 0.13 |
| HMGB1 | 101 | 13 | 0 | 0 | ng/ml | 2.19 | 0.72 | 0.33 | 2.19 | 0.72 | 0.33 | 2.21 | 0.70 | 0.32 |
| ICAM1 | 112 | 2 | 0 | 0 | pg/ml | 10.0 | 0.82 | 0.082 | 10.1 | 0.72 | 0.07 | 10.1 | 0.72 | 0.07 |
| IFNγ | 112 | 2 | 0 | 0 | pg/ml | 4.66 | 0.46 | 0.099 | 4.66 | 0.46 | 0.10 | 4.67 | 0.46 | 0.10 |
| IL1β | 112 | 2 | 0 | 0 | pg/ml | 7.72 | 1.48 | 0.19 | 7.72 | 1.48 | 0.19 | 7.68 | 1.49 | 0.19 |
| IL5 | 112 | 2 | 0 | 0 | pg/ml | 2.94 | 0.40 | 0.14 | 2.94 | 0.40 | 0.14 | 2.95 | 0.40 | 0.13 |
| IL6 | 112 | 2 | 0 | 0 | pg/ml | 3.15 | 0.79 | 0.25 | 3.15 | 0.79 | 0.25 | 3.15 | 0.78 | 0.25 |
| IL8 | 112 | 2 | 0 | 0 | pg/ml | 8.66 | 0.51 | 0.06 | 8.66 | 0.51 | 0.06 | 8.66 | 0.51 | 0.06 |
| IL10 | 111 | 3 | 1 | 0 | pg/ml | 1.40 | 0.93 | 0.67 | 1.43 | 0.77 | 0.54 | 1.45 | 0.77 | 0.53 |
| IL17A | 112 | 2 | 0 | 0 | pg/ml | 5.01 | 0.67 | 0.13 | 5.01 | 0.67 | 0.13 | 5.01 | 0.67 | 0.13 |
| MMP9 | 112 | 2 | 1 | 0 | pg/ml | 14.4 | 1.32 | 0.09 | 14.4 | 1.32 | 0.09 | 14.4 | 1.31 | 0.09 |
| MPO | 113 | 1 | 0 | 0 | pg/ml | 15.8 | 1.27 | 0.08 | 15.8 | 1.24 | 0.08 | 15.8 | 1.24 | 0.08 |
| NE | 114 | 0 | 8 | 0 | µg/ml | 2.80 | 1.33 | 0.47 | 2.60 | 1.48 | 0.57 | 2.60 | 1.48 | 0.57 |
| PR3§ | 113 | 1 | 0 | 22 | pg/ml | 14.4 | 1.22 | 0.08 | 14.7 | 1.22 | 0.08 | 14.7 | 1.21 | 0.08 |
| SRAGE | 112 | 2 | 13 | 0 | pg/ml | 6.38 | 1.08 | 0.17 | 6.07 | 1.34 | 0.22 | 6.05 | 1.35 | 0.22 |
| S100A8 | 112 | 2 | 0 | 0 | pg/ml | 7.79 | 0.80 | 0.10 | 7.79 | 0.80 | 0.10 | 7.81 | 0.8 | 0.10 |
| S100A9 | 112 | 2 | 0 | 2 | pg/ml | 8.99 | 0.95 | 0.11 | 9.04 | 1.03 | 0.11 | 9.02 | 1.03 | 0.11 |
| SLPI | 112 | 2 | 0 | 0 | pg/ml | 11.7 | 1.51 | 0.13 | 11.7 | 1.51 | 0.13 | 11.7 | 1.50 | 0.13 |
| TARC | 110 | 4 | 2 | 0 | pg/ml | 5.72 | 0.27 | 0.05 | 5.72 | 0.27 | 0.05 | 5.73 | 0.27 | 0.05 |
| TNFα | 112 | 2 | 0 | 0 | pg/ml | 4.84 | 0.89 | 0.18 | 4.84 | 0.89 | 0.18 | 4.85 | 0.89 | 0.18 |
| YKL40 | 112 | 2 | 0 | 9 | pg/ml | 12.0 | 1.08 | 0.09 | 12.1 | 1.11 | 0.09 | 12.1 | 1.10 | 0.09 |

* Additional information on Raw (Unadjusted) and Imputed Biomarker Measurements to accompany main text Table 2.

† **Abbreviations**: **CRP** C reactive protein; **ENRAGE** Extracellular newly identified receptor for advanced glycation end products binding protein; **GMCSF** granulocyte macrophage colony stimulating factor; **HMGB1** high mobility group box 1 protein; **ICAM1** intracellular adhesion molecule 1; **IFNγ** interferon γ; **IL** interleukin; **MMP9** matrix metallopeptidase 9; **MPO** myeloperoxidase; **NE** neutrophil elastase; **PR3** proteinase 3; **sRAGE** soluble receptor for advanced glycation products; **SLPI** secretory leukoprotease inhibitor; **TARC** thymus and activation regulation chemokine; **TNFα** tumor necrosis factor α; **YKL40** chitinase 3-like 1 protein; **SD** Standard Deviation; **CV** Coefficient of Variation.

§ After log transformation, adjusted and imputed data for PR-3 were greater than raw data, *t*-test $p = 0.001$, both comparisons.
