## Supplemental Table 5 for "Associations of Sputum Biomarkers with Clinical Outcomes in People with Cystic Fibrosis"

**Supplemental Table 5. Pearson Correlations between Biomarkers.**

| | Calprotectin | CRP | ENRAGE | GMCSF | HMGB1 | ICAM1 | IFNG | IL1B | IL5 | IL6 | IL8 |
|---|---|---|---|---|---|---|---|---|---|---|---|
| Calprotectin | | | | | | | | 0.41 | | | |
| CRP | | | | -0.42 | | | | | | | |
| ENRAGE | | | | 0.60 | | 0.47 | | 0.53 | | | 0.43 |
| GMCSF | | -0.42 | 0.60 | | | 0.49 | 0.57 | | 0.62 | | 0.45 |
| HMGB1 | | | | | | | | | | | |
| ICAM1 | | | 0.47 | 0.49 | | | 0.48 | 0.47 | 0.41 | 0.48 | |
| IFNγ | | | | 0.57 | | 0.48 | | | 0.91 | 0.41 | |
| IL1β | 0.41 | | 0.53 | | | 0.47 | | | | | 0.67 |
| IL5 | | | | 0.62 | | 0.41 | 0.91 | | | | |
| IL6 | | | | | | 0.48 | 0.41 | | | | |
| IL8 | | | 0.43 | 0.45 | | | | 0.67 | | | |
| IL10 | | | | 0.44 | | | 0.74 | | 0.65 | 0.55 | |
| IL17A | | | | 0.52 | | 0.42 | 0.78 | | 0.76 | | |
| MMP9 | | | 0.72 | 0.46 | | 0.54 | | 0.86 | | | 0.62 |
| MPO | | | 0.88 | 0.54 | | 0.50 | | 0.46 | | | 0.50 |
| NE | 0.45 | | | | | 0.42 | | 0.79 | | | 0.51 |
| PR3 | 0.41 | | | | | | | 0.75 | | | 0.69 |
| sRAGE | | | 0.45 | | | 0.53 | 0.50 | 0.69 | 0.54 | | |
| S100A8 | | | | | | | | | | 0.43 | |
| S100A9 | | | | | | | | -0.43 | | | |
| SLPI | | | | | | | | -0.47 | | 0.57 | |
| TARC | | | | 0.46 | | | 0.91 | | 0.89 | | |
| TNFα | | | | 0.42 | | | 0.53 | | 0.43 | | |
| YKL40 | | | | | | 0.54 | | 0.76 | | | 0.66 |

| | IL17A | MMP9 | MPO | NE | PR3 | sRAGE | S100A8 | S100A9 | SLPI | TARC | TNFA | YKL40 |
|---|---|---|---|---|---|---|---|---|---|---|---|---|
| Calprotectin | | | | 0.45 | 0.41 | | | | | | | |
| CRP | | | | | | | | | | | | |
| ENRAGE | | 0.72 | 0.88 | | | 0.45 | | | | | | |
| GMCSF | 0.52 | 0.46 | 0.54 | | | | | | | 0.46 | 0.42 | |
| HMGB1 | | | | | | | | | | | | |
| ICAM1 | 0.42 | 0.54 | 0.50 | 0.42 | | 0.53 | | | | | | 0.54 |
| IFNγ | 0.78 | | | | | 0.50 | | | | 0.91 | 0.53 | |
| IL1β | | 0.86 | 0.46 | 0.79 | 0.75 | 0.69 | | -0.43 | -0.47 | | | 0.76 |
| IL5 | 0.76 | | | | | 0.54 | | | | 0.89 | 0.43 | |
| IL6 | | | | | | | 0.43 | | 0.57 | | | |
| IL8 | | 0.62 | 0.50 | 0.51 | 0.69 | | | | | | | 0.66 |
| IL10 | 0.61 | | | | | | | | 0.41 | 0.65 | 0.45 | |
| IL17A | | | | | | 0.60 | | | | 0.71 | 0.65 | |
| MMP9 | | | 0.64 | 0.68 | 0.65 | 0.65 | | -0.42 | -0.44 | | | 0.66 |
| MPO | | 0.64 | | 0.46 | | 0.48 | | | | | | 0.43 |
| NE | | 0.68 | 0.46 | | 0.73 | 0.80 | -0.44 | -0.44 | -0.60 | | | 0.79 |
| PR3 | | 0.65 | | 0.73 | | 0.51 | | | -0.43 | | | 0.73 |
| sRAGE | 0.60 | 0.65 | 0.48 | 0.80 | 0.51 | | | | -0.48 | 0.49 | | 0.69 |
| S100A8 | | | | -0.44 | | | | | | | | |
| S100A9 | | -0.42 | | -0.44 | | | | | | | | -0.45 |
| SLPI | | -0.44 | | -0.60 | -0.43 | -0.48 | | | | | | |
| TARC | 0.71 | | | | | 0.49 | | | | | 0.47 | |
| TNFα | 0.65 | | | | | | | | | 0.47 | | |
| YKL40 | | 0.66 | 0.43 | 0.79 | 0.73 | 0.69 | | -0.45 | | | | |

* All positive and negative correlations shown were significant after stringent Bonferroni correction; all have uncorrected $p < 1\times10^{-5}$. Correlations were identical whether using data only adjusted for biomarker detection limits or adjusted and imputed for missingness.

† **Abbreviations**: **CRP** C reactive protein; **ENRAGE** Extracellular newly identified receptor for advanced glycation end products binding protein; **GMCSF** granulocyte macrophage colony stimulating factor; **HMGB1** high mobility group box 1 protein; **ICAM1** intracellular adhesion molecule 1; **IFNγ** interferon γ; **IL** interleukin; **MMP9** matrix metallopeptidase 9; **MPO** myeloperoxidase; **NE** neutrophil elastase; **PR3** proteinase 3; **sRAGE** soluble receptor for advanced glycation products; **SLPI** secretory leukoprotease inhibitor; **TARC** thymus and activation regulation chemokine; **TNFα** tumor necrosis factor α; **YKL40** chitinase 3-like 1 protein.
