## Supplemental Table 6 for "Associations of Sputum Biomarkers with Clinical Outcomes in People with Cystic Fibrosis"

**Supplemental Table 6. Univariable Proportional Hazards Models of Time to Next Pulmonary Exacerbation**

| Biomarker* | Data Adjusted for Detection Limits | | | | | Data Adjusted and Imputed for Missingness | | | | |
|---|---|---|---|---|---|---|---|---|---|---|
| | **Estimate** | **SE** | **Hazard Ratio** | ***p*** | **95% CI of the Hazard Ratio** | **Estimate** | **SE** | **Hazard Ratio** | ***p*** | **95% CI of the Hazard Ratio** |
| MPO | 0.24 | 0.10 | 1.28 | 0.010 | 1.06 to 1.54 | 0.24 | 0.09 | 1.27 | 0.010 | 1.06 to 1.53 |
| NE | 0.18 | 0.08 | 1.20 | 0.018 | 1.03 to 1.39 | 0.18 | 0.08 | 1.20 | 0.018 | 1.03 to 1.39 |
| S100A9 | -0.27 | 0.12 | 0.76 | 0.028 | 0.60 to 0.97 | -0.19 | 0.13 | 0.82 | 0.130 | 0.64 to 1.06 |
| SRAGE | 0.20 | 0.09 | 1.22 | 0.030 | 1.02 to 1.45 | 0.21 | 0.09 | 1.24 | 0.020 | 1.03 to 1.48 |
| ENRAGE | 0.20 | 0.09 | 1.22 | 0.032 | 1.02 to 1.46 | 0.20 | 0.09 | 1.22 | 0.029 | 1.02 to 1.46 |
| IL1β | 0.16 | 0.08 | 1.17 | 0.041 | 1.01 to 1.37 | 0.16 | 0.08 | 1.17 | 0.039 | 1.01 to 1.37 |
| YKL40 | 0.19 | 0.10 | 1.21 | 0.055 | 1.00 to 1.46 | 0.20 | 0.09 | 1.22 | 0.036 | 1.01 to 1.46 |
| IL17A | 0.30 | 0.17 | 1.35 | 0.078 | 0.97 to 1.88 | 0.29 | 0.17 | 1.33 | 0.084 | 0.96 to 1.84 |
| PR3 | 0.14 | 0.09 | 1.15 | 0.110 | 0.97 to 1.37 | 0.17 | 0.10 | 1.18 | 0.086 | 0.98 to 1.43 |
| ICAM1 | 0.24 | 0.15 | 1.27 | 0.120 | 0.94 to 1.71 | 0.22 | 0.14 | 1.25 | 0.100 | 0.96 to 1.64 |
| IL8 | 0.36 | 0.24 | 1.44 | 0.130 | 0.90 to 2.29 | 0.35 | 0.23 | 1.43 | 0.130 | 0.90 to 2.26 |
| Calprotectin | 0.05 | 0.04 | 1.06 | 0.130 | 0.98 to 1.13 | 0.05 | 0.04 | 1.06 | 0.130 | 0.98 to 1.13 |
| MMP9 | 0.13 | 0.08 | 1.13 | 0.130 | 0.96 to 1.34 | 0.16 | 0.08 | 1.18 | 0.050 | 1.00 to 1.38 |
| SLPI | -0.09 | 0.08 | 0.91 | 0.250 | 0.78 to 1.07 | -0.10 | 0.08 | 0.91 | 0.240 | 0.78 to 1.06 |
| IL5 | 0.32 | 0.30 | 1.38 | 0.290 | 0.76 to 2.51 | 0.30 | 0.30 | 1.35 | 0.320 | 0.75 to 2.43 |
| S100A8 | -0.15 | 0.14 | 0.86 | 0.300 | 0.65 to 1.14 | -0.13 | 0.14 | 0.88 | 0.340 | 0.66 to 1.15 |
| TARC | 0.40 | 0.44 | 1.49 | 0.370 | 0.62 to 3.55 | 0.33 | 0.42 | 1.39 | 0.430 | 0.61 to 3.13 |
| GMCSF | 0.24 | 0.27 | 1.27 | 0.370 | 0.75 to 2.15 | 0.15 | 0.31 | 1.16 | 0.640 | 0.63 to 2.13 |
| HMGB1 | 0.11 | 0.15 | 1.12 | 0.450 | 0.84 to 1.49 | 0.11 | 0.15 | 1.11 | 0.470 | 0.84 to 1.48 |
| TNFα | -0.08 | 0.12 | 0.93 | 0.510 | 0.73 to 1.17 | -0.09 | 0.12 | 0.92 | 0.460 | 0.73 to 1.16 |
| IFNγ | 0.15 | 0.25 | 1.16 | 0.560 | 0.71 to 1.9 | 0.13 | 0.25 | 1.14 | 0.600 | 0.70 to 1.86 |
| IL6 | -0.05 | 0.14 | 0.95 | 0.720 | 0.72 to 1.26 | -0.04 | 0.14 | 0.96 | 0.760 | 0.73 to 1.26 |
| IL10 | -0.03 | 0.15 | 0.97 | 0.830 | 0.72 to 1.29 | -0.03 | 0.15 | 0.97 | 0.830 | 0.72 to 1.3 |
| CRP | 0.02 | 0.18 | 1.02 | 0.890 | 0.72 to 1.46 | 0.05 | 0.34 | 1.05 | 0.890 | 0.54 to 2.05 |

* **Abbreviations**: **CRP** C reactive protein; **ENRAGE** Extracellular newly identified receptor for advanced glycation end products binding protein; **GMCSF** granulocyte macrophage colony stimulating factor; **HMGB1** high mobility group box 1 protein; **ICAM1** intracellular adhesion molecule 1; **IFNγ** interferon γ; **IL** interleukin; **MMP9** matrix metallopeptidase 9; **MPO** myeloperoxidase; **NE** neutrophil elastase; **PR3** proteinase 3; **sRAGE** soluble receptor for advanced glycation products; **SLPI** secretory leukoprotease inhibitor; **TARC** thymus and activation regulation chemokine; **TNFα** tumor necrosis factor α; **YKL40** chitinase 3-like 1 protein.
