## Supplemental Table 7 for "Associations of Sputum Biomarkers with Clinical Outcomes in People with Cystic Fibrosis"

**Supplemental Table 7A. Univariable Associations of Age with Biomarkers**

| Univariable Biomarker* | Data Adjusted for Detection Limits | | | | Data Adjusted and Imputed for Missingness | | | |
|---|---|---|---|---|---|---|---|---|
| | **Estimate** | **SE** | ***p*** | **95% CI** | **Estimate** | **SE** | ***p*** | **95% CI** |
| MPO | 0.96 | 0.92 | 0.30 | -0.84 to 2.75 | 1.10 | 0.90 | 0.21 | -0.64 to 2.89 |
| NE | 1.80 | 0.75 | 0.02 | 0.37 to 3.33 | 1.80 | 0.75 | 0.02 | 0.37 to 3.33 |
| S100A9 | 0.22 | 1.10 | 0.85 | -1.96 to 2.39 | 0.63 | 1.20 | 0.60 | -1.73 to 2.98 |
| SRAGE | 2.00 | 0.84 | 0.02 | 0.33 to 3.61 | 2.10 | 0.84 | 0.02 | 0.43 to 3.70 |
| ENRAGE | 1.60 | 0.93 | 0.09 | -0.24 to 3.40 | 1.70 | 0.94 | 0.08 | -0.19 to 3.49 |
| IL1β | 0.98 | 0.77 | 0.20 | -0.53 to 2.49 | 1.10 | 0.77 | 0.16 | -0.41 to 2.61 |
| YKL40 | 1.20 | 1.00 | 0.26 | -0.86 to 3.18 | 0.66 | 1.00 | 0.52 | -1.35 to 2.66 |
| IL17A | 0.74 | 1.70 | 0.67 | -2.61 to 4.09 | 0.76 | 1.70 | 0.65 | -2.55 to 4.07 |
| PR3 | 0.59 | 0.93 | 0.53 | -1.24 to 2.42 | 1.20 | 0.99 | 0.25 | -0.79 to 3.09 |
| ICAM1 | -0.57 | 1.60 | 0.72 | -3.69 to 2.54 | 0.48 | 1.40 | 0.73 | -2.23 to 3.19 |
| IL8 | 2.10 | 2.20 | 0.36 | -2.33 to 6.49 | 2.10 | 2.30 | 0.36 | -2.37 to 6.50 |
| Calprotectin | -0.17 | 0.39 | 0.65 | -0.93 to 0.59 | -0.17 | 0.39 | 0.66 | -0.93 to 0.59 |
| MMP9 | 1.40 | 0.86 | 0.10 | -0.24 to 3.12 | 1.50 | 0.86 | 0.09 | -0.21 to 3.19 |
| SLPI | -2.01 | 0.73 | 0.01 | -3.45 to -0.57 | -2.10 | 0.73 | 0.004 | -3.58 to -0.72 |
| IL5 | 3.70 | 2.80 | 0.20 | -1.91 to 9.23 | 2.80 | 2.80 | 0.33 | -2.73 to 8.25 |
| S100A8 | -1.00 | 1.40 | 0.48 | -3.84 to 1.80 | -1.30 | 1.40 | 0.34 | -4.10 to 1.42 |
| TARC | 6.50 | 4.20 | 0.13 | -1.81 to 14.7 | 5.60 | 4.10 | 0.18 | -2.54 to 13.7 |
| GMCSF | 1.30 | 3.10 | 0.68 | -4.85 to 7.41 | 2.20 | 3.50 | 0.54 | -4.75 to 9.16 |
| HMGB1 | -0.92 | 1.70 | 0.60 | -4.31 to 2.47 | -1.00 | 1.50 | 0.49 | -3.89 to 1.85 |
| TNFα | 0.67 | 1.30 | 0.60 | -1.84 to 3.18 | 0.12 | 1.30 | 0.92 | -2.38 to 2.62 |
| IFNγ | 3.90 | 2.50 | 0.12 | -0.96 to 8.72 | 3.70 | 2.50 | 0.14 | -1.17 to 8.55 |
| IL6 | -1.00 | 1.40 | 0.47 | -3.88 to 1.80 | -1.20 | 1.50 | 0.42 | -4.04 to 1.67 |
| IL10 | -0.64 | 1.50 | 0.67 | -3.60 to 2.32 | -0.61 | 1.60 | 0.70 | -3.69 to 2.47 |
| CRP | 1.30 | 1.90 | 0.49 | -2.40 to 5.04 | 2.30 | 3.40 | 0.51 | -4.47 to 9.01 |

* **Abbreviations**: **CRP** C reactive protein; **ENRAGE** Extracellular newly identified receptor for advanced glycation end products binding protein; **GMCSF** granulocyte macrophage colony stimulating factor; **HMGB1** high mobility group box 1 protein; **ICAM1** intracellular adhesion molecule 1; **IFNγ** interferon γ; **IL** interleukin; **MMP9** matrix metallopeptidase 9; **MPO** myeloperoxidase; **NE** neutrophil elastase; **PR3** proteinase 3; **sRAGE** soluble receptor for advanced glycation products; **SLPI** secretory leukoprotease inhibitor; **TARC** thymus and activation regulation chemokine; **TNFα** tumor necrosis factor α; **YKL40** chitinase 3-like 1 protein.

**Supplemental Table 7B. Univariable Associations of Weight-for-Age *z*-Score with Biomarkers**

| Univariable Biomarker* | Data Adjusted for Detection Limits | | | | Data Adjusted and Imputed for Missingness | | | |
|---|---|---|---|---|---|---|---|---|
| | **Estimate** | **SE** | ***p*** | **95% CI** | **Estimate** | **SE** | ***p*** | **95% CI** |
| MPO | 0.04 | 0.07 | 0.62 | -0.11 to 0.18 | 0.05 | 0.07 | 0.48 | -0.09 to 0.19 |
| NE | -0.06 | 0.06 | 0.33 | -0.18 to 0.06 | -0.06 | 0.06 | 0.33 | -0.18 to 0.06 |
| S100A9 | 0.01 | 0.09 | 0.95 | -0.17 to 0.18 | 0.04 | 0.10 | 0.67 | -0.15 to 0.23 |
| SRAGE | -0.04 | 0.07 | 0.60 | -0.17 to 0.10 | -0.04 | 0.07 | 0.6 | -0.17 to 0.10 |
| ENRAGE | 0.07 | 0.08 | 0.35 | -0.08 to 0.22 | 0.08 | 0.08 | 0.32 | -0.07 to 0.23 |
| IL1β | -0.02 | 0.06 | 0.71 | -0.15 to 0.10 | -0.01 | 0.06 | 0.87 | -0.13 to 0.11 |
| YKL40 | -0.01 | 0.08 | 0.95 | -0.17 to 0.16 | 0.04 | 0.08 | 0.65 | -0.12 to 0.20 |
| IL17A | 0.21 | 0.14 | 0.12 | -0.05 to 0.48 | 0.25 | 0.13 | 0.067 | -0.01 to 0.51 |
| PR3 | -0.06 | 0.08 | 0.41 | -0.21 to 0.08 | 0.03 | 0.08 | 0.75 | -0.13 to 0.18 |
| ICAM1 | 0.26 | 0.13 | 0.04 | 0.01 to 0.50 | 0.26 | 0.11 | 0.017 | 0.05 to 0.48 |
| IL8 | 0.12 | 0.18 | 0.50 | -0.23 to 0.48 | 0.14 | 0.18 | 0.45 | -0.22 to 0.50 |
| Calprotectin | -0.02 | 0.03 | 0.50 | -0.08 to 0.04 | -0.02 | 0.03 | 0.5 | -0.08 to 0.04 |
| MMP9 | 0.03 | 0.07 | 0.65 | -0.11 to 0.17 | 0.04 | 0.07 | 0.61 | -0.10 to 0.17 |
| SLPI | 0.06 | 0.06 | 0.35 | -0.06 to 0.18 | 0.05 | 0.06 | 0.41 | -0.07 to 0.17 |
| IL5 | 0.34 | 0.23 | 0.14 | -0.10 to 0.79 | 0.33 | 0.22 | 0.14 | -0.11 to 0.78 |
| S100A8 | 0.43 | 0.11 | <0.001 | 0.21 to 0.64 | 0.41 | 0.11 | <0.001 | 0.20 to 0.62 |
| TARC | 0.34 | 0.34 | 0.33 | -0.34 to 1.01 | 0.37 | 0.34 | 0.27 | -0.29 to 1.03 |
| GMCSF | 0.65 | 0.24 | 0.009 | 0.17 to 1.13 | 0.83 | 0.28 | 0.0034 | 0.28 to 1.37 |
| HMGB1 | 0.01 | 0.14 | 0.96 | -0.27 to 0.28 | 0.05 | 0.12 | 0.65 | -0.18 to 0.29 |
| TNFα | 0.17 | 0.10 | 0.10 | -0.03 to 0.37 | 0.13 | 0.10 | 0.19 | -0.06 to 0.34 |
| IFNγ | 0.43 | 0.20 | 0.03 | 0.048 to 0.82 | 0.40 | 0.20 | 0.045 | 0.01 to 0.79 |
| IL6 | 0.30 | 0.11 | 0.008 | 0.082 to 0.53 | 0.30 | 0.11 | 0.0094 | 0.08 to 0.53 |
| IL10 | 0.33 | 0.12 | 0.007 | 0.094 to 0.56 | 0.36 | 0.12 | 0.0035 | 0.12 to 0.60 |
| CRP | 0.06 | 0.15 | 0.67 | -0.24 to 0.37 | -0.14 | 0.28 | 0.62 | -0.68 to 0.40 |

* **Abbreviations**: **CRP** C reactive protein; **ENRAGE** Extracellular newly identified receptor for advanced glycation end products binding protein; **GMCSF** granulocyte macrophage colony stimulating factor; **HMGB1** high mobility group box 1 protein; **ICAM1** intracellular adhesion molecule 1; **IFNγ** interferon γ; **IL** interleukin; **MMP9** matrix metallopeptidase 9; **MPO** myeloperoxidase; **NE** neutrophil elastase; **PR3** proteinase 3; **sRAGE** soluble receptor for advanced glycation products; **SLPI** secretory leukoprotease inhibitor; **TARC** thymus and activation regulation chemokine; **TNFα** tumor necrosis factor α; **YKL40** chitinase 3-like 1 protein.

**Supplemental Table 7C. Univariable Associations of Diabetes with Biomarkers**

| Univariable Biomarker* | Data Adjusted for Detection Limits | | | | Data Adjusted and Imputed for Missingness | | | |
|---|---|---|---|---|---|---|---|---|
| | **Estimate** | **SE** | ***p*** | **95% CI** | **Estimate** | **SE** | ***p*** | **95% CI** |
| MPO | 0.05 | 0.03 | 0.13 | -0.01 to 0.11 | 0.05 | 0.03 | 0.14 | -0.02 to 0.11 |
| NE | 0.02 | 0.03 | 0.42 | -0.03 to 0.07 | 0.02 | 0.03 | 0.42 | -0.03 to 0.07 |
| S100A9 | -0.04 | 0.04 | 0.26 | -0.12 to 0.03 | -0.04 | 0.04 | 0.31 | -0.12 to 0.04 |
| SRAGE | -0.03 | 0.03 | 0.26 | -0.09 to 0.02 | -0.02 | 0.03 | 0.41 | -0.08 to 0.03 |
| ENRAGE | 0.02 | 0.03 | 0.46 | -0.04 to 0.09 | 0.03 | 0.03 | 0.42 | -0.04 to 0.09 |
| IL1β | -0.01 | 0.03 | 0.79 | -0.06 to 0.04 | -0.01 | 0.03 | 0.85 | -0.06 to 0.05 |
| YKL40 | 0.02 | 0.04 | 0.60 | -0.04 to 0.09 | 0.03 | 0.04 | 0.45 | -0.04 to 0.10 |
| IL17A | -0.13 | 0.06 | 0.02 | -0.24 to -0.02 | -0.11 | 0.06 | 0.05 | -0.22 to 0.00 |
| PR3 | 0.01 | 0.03 | 0.65 | -0.05 to 0.08 | 0.00 | 0.03 | 0.90 | -0.06 to 0.07 |
| ICAM1 | -0.04 | 0.05 | 0.47 | -0.14 to 0.07 | -0.02 | 0.05 | 0.73 | -0.11 to 0.08 |
| IL8 | 0.06 | 0.08 | 0.43 | -0.09 to 0.21 | 0.07 | 0.08 | 0.40 | -0.09 to 0.22 |
| Calprotectin | -0.002 | 0.01 | 0.90 | -0.03 to 0.02 | -0.00 | 0.01 | 0.91 | -0.03 to 0.02 |
| MMP9 | 0.01 | 0.03 | 0.85 | -0.05 to 0.06 | 0.02 | 0.03 | 0.56 | -0.04 to 0.07 |
| SLPI | -0.03 | 0.02 | 0.26 | -0.08 to 0.02 | -0.02 | 0.03 | 0.34 | -0.08 to 0.03 |
| IL5 | -0.18 | 0.10 | 0.07 | -0.36 to 0.01 | -0.19 | 0.10 | 0.06 | -0.38 to 0.00 |
| S100A8 | 0.02 | 0.05 | 0.74 | -0.08 to 0.11 | 0.04 | 0.05 | 0.43 | -0.06 to 0.13 |
| TARC | -0.31 | 0.14 | 0.03 | -0.58 to -0.04 | -0.37 | 0.14 | 0.01 | -0.64 to -0.10 |
| GMCSF | -0.08 | 0.11 | 0.47 | -0.28 to 0.13 | -0.08 | 0.12 | 0.51 | -0.32 to 0.16 |
| HMGB1 | -0.02 | 0.06 | 0.79 | -0.12 to 0.09 | -0.01 | 0.06 | 0.80 | -0.12 to 0.10 |
| TNFα | -0.07 | 0.04 | 0.11 | -0.15 to 0.02 | -0.081 | 0.04 | 0.07 | -0.17 to 0.00 |
| IFNγ | -0.19 | 0.08 | 0.02 | -0.35 to -0.03 | -0.20 | 0.08 | 0.02 | -0.36 to -0.04 |
| IL6 | -0.06 | 0.05 | 0.21 | -0.16 to 0.03 | -0.06 | 0.05 | 0.20 | -0.16 to 0.03 |
| IL10 | -0.15 | 0.05 | 0.002 | -0.25 to -0.06 | -0.16 | 0.05 | 0.002 | -0.26 to -0.06 |
| CRP | 0.04 | 0.06 | 0.58 | -0.09 to 0.16 | 0.11 | 0.12 | 0.37 | -0.12 to 0.34 |

* **Abbreviations**: **CRP** C reactive protein; **ENRAGE** Extracellular newly identified receptor for advanced glycation end products binding protein; **GMCSF** granulocyte macrophage colony stimulating factor; **HMGB1** high mobility group box 1 protein; **ICAM1** intracellular adhesion molecule 1; **IFNγ** interferon γ; **IL** interleukin; **MMP9** matrix metallopeptidase 9; **MPO** myeloperoxidase; **NE** neutrophil elastase; **PR3** proteinase 3; **sRAGE** soluble receptor for advanced glycation products; **SLPI** secretory leukoprotease inhibitor; **TARC** thymus and activation regulation chemokine; **TNFα** tumor necrosis factor α; **YKL40** chitinase 3-like 1 protein.

**Supplemental Table 7D. Univariable Associations of Prior Pulmonary Exacerbations with Biomarkers**

| Univariable Biomarker* | Data Adjusted for Detection Limits | | | | Data Adjusted and Imputed for Missingness | | | |
|---|---|---|---|---|---|---|---|---|
| | **Estimate** | **SE** | ***p*** | **95% CI** | **Estimate** | **SE** | ***p*** | **95% CI** |
| MPO | 0.20 | 0.12 | 0.09 | -0.03 to 0.44 | 0.21 | 0.12 | 0.08 | -0.02 to 0.44 |
| NE | 0.20 | 0.10 | 0.05 | 0.00 to 0.39 | 0.20 | 0.10 | 0.05 | 0.00 to 0.39 |
| S100A9 | -0.26 | 0.15 | 0.08 | -0.54 to 0.03 | -0.26 | 0.16 | 0.10 | -0.57 to 0.04 |
| SRAGE | 0.16 | 0.11 | 0.15 | -0.06 to 0.38 | 0.16 | 0.11 | 0.16 | -0.06 to 0.37 |
| ENRAGE | 0.12 | 0.12 | 0.35 | -0.13 to 0.36 | 0.11 | 0.12 | 0.36 | -0.13 to 0.36 |
| IL1β | 0.16 | 0.10 | 0.12 | -0.04 to 0.36 | 0.16 | 0.10 | 0.12 | -0.04 to 0.36 |
| YKL40 | 0.08 | 0.14 | 0.57 | -0.19 to 0.35 | 0.04 | 0.13 | 0.79 | -0.23 to 0.30 |
| IL17A | 0.15 | 0.23 | 0.50 | -0.29 to 0.60 | 0.11 | 0.22 | 0.61 | -0.32 to 0.55 |
| PR3 | 0.24 | 0.12 | 0.06 | -0.00 to 0.48 | 0.22 | 0.13 | 0.09 | -0.04 to 0.47 |
| ICAM1 | 0.07 | 0.21 | 0.73 | -0.34 to 0.48 | 0.04 | 0.18 | 0.81 | -0.32 to 0.40 |
| IL8 | 0.48 | 0.30 | 0.11 | -0.10 to 1.06 | 0.47 | 0.29 | 0.12 | -0.11 to 1.04 |
| Calprotectin | 0.13 | 0.05 | 0.01 | 0.03 to 0.22 | 0.13 | 0.05 | 0.01 | 0.03 to 0.22 |
| MMP9 | 0.12 | 0.11 | 0.29 | -0.10 to 0.35 | 0.13 | 0.11 | 0.23 | -0.08 to 0.35 |
| SLPI | -0.18 | 0.10 | 0.07 | -0.38 to 0.01 | -0.19 | 0.10 | 0.06 | -0.38 to 0.00 |
| IL5 | -0.16 | 0.38 | 0.68 | -0.90 to 0.58 | -0.14 | 0.37 | 0.70 | -0.88 to 0.59 |
| S100A8 | -0.26 | 0.19 | 0.17 | -0.63 to 0.11 | -0.29 | 0.18 | 0.12 | -0.65 to 0.07 |
| TARC | -0.96 | 0.56 | 0.09 | -2.05 to 0.14 | -0.73 | 0.53 | 0.17 | -1.77 to 0.32 |
| GMCSF | -0.00 | 0.41 | 1.00 | -0.81 to 0.81 | -0.11 | 0.46 | 0.82 | -1.02 to 0.80 |
| HMGB1 | 0.47 | 0.22 | 0.03 | 0.04 to 0.90 | 0.51 | 0.21 | 0.02 | 0.10 to 0.92 |
| TNFα | -0.20 | 0.17 | 0.24 | -0.53 to 0.13 | -0.18 | 0.17 | 0.28 | -0.51 to 0.15 |
| IFNγ | -0.15 | 0.33 | 0.64 | -0.80 to 0.49 | -0.15 | 0.33 | 0.65 | -0.79 to 0.50 |
| IL6 | -0.26 | 0.19 | 0.18 | -0.63 to 0.12 | -0.25 | 0.19 | 0.20 | -0.62 to 0.13 |
| IL10 | -0.41 | 0.20 | 0.04 | -0.80 to -0.03 | -0.40 | 0.20 | 0.05 | -0.80 to -0.01 |
| CRP | 0.32 | 0.25 | 0.20 | -0.17 to 0.80 | 0.18 | 0.45 | 0.69 | -0.70 to 1.06 |

* **Abbreviations**: **CRP** C reactive protein; **ENRAGE** Extracellular newly identified receptor for advanced glycation end products binding protein; **GMCSF** granulocyte macrophage colony stimulating factor; **HMGB1** high mobility group box 1 protein; **ICAM1** intracellular adhesion molecule 1; **IFNγ** interferon γ; **IL** interleukin; **MMP9** matrix metallopeptidase 9; **MPO** myeloperoxidase; **NE** neutrophil elastase; **PR3** proteinase 3; **sRAGE** soluble receptor for advanced glycation products; **SLPI** secretory leukoprotease inhibitor; **TARC** thymus and activation regulation chemokine; **TNFα** tumor necrosis factor α; **YKL40** chitinase 3-like 1 protein.

**Supplemental Table 7E. Univariable Associations of $FEV_1$% with Biomarkers**

| Univariable Biomarker* | Data Adjusted for Detection Limits | | | | Data Adjusted and Imputed for Missingness | | | |
|---|---|---|---|---|---|---|---|---|
| | Estimate† | SE | *p* | 95% CI | Estimate† | SE | *p* | 95% CI |
| MPO§ | -6.30 | 1.60 | <0.001 | -9.38 to -3.27 | -6.20 | 1.50 | <0.001 | -9.18 to -3.14 |
| NE§ | -5.60 | 1.30 | <0.001 | -8.17 to -3.09 | -5.60 | 1.30 | <0.001 | -8.17 to -3.09 |
| S100A9 | 3.10 | 2.00 | 0.13 | -0.85 to 6.98 | 1.30 | 2.20 | 0.54 | -2.94 to 5.58 |
| SRAGE¶ | -5.20 | 1.50 | <0.001 | -8.12 to -2.36 | -5.50 | 1.50 | <0.001 | -8.36 to -2.64 |
| ENRAGE¶ | -5.30 | 1.60 | 0.001 | -8.53 to -2.12 | -5.40 | 1.60 | 0.001 | -8.67 to -2.22 |
| IL1β | -3.70 | 1.40 | 0.007 | -6.41 to -1.07 | -3.80 | 1.40 | 0.006 | -6.48 to -1.11 |
| YKL40 | -3.60 | 1.90 | 0.06 | -7.19 to 0.08 | -2.10 | 1.90 | 0.27 | -5.69 to 1.57 |
| IL17A | -0.88 | 3.10 | 0.78 | -6.96 to 5.21 | -1.10 | 3.10 | 0.71 | -7.18 to 4.91 |
| PR3 | -4.10 | 1.70 | 0.01 | -7.35 to -0.87 | -5.40 | 1.70 | 0.002 | -8.80 to -1.97 |
| ICAM1 | -0.72 | 2.90 | 0.80 | -6.37 to 4.92 | -1.70 | 2.50 | 0.49 | -6.71 to 3.23 |
| IL8 | -8.00 | 4.00 | 0.05 | -15.9 to -0.10 | -8.00 | 4.00 | 0.051 | -15.9 to -0.05 |
| Calprotectin | -1.60 | 0.69 | 0.02 | -2.91 to -0.22 | -1.60 | 0.69 | 0.024 | -2.91 to -0.22 |
| MMP9¶ | -5.00 | 1.50 | 0.001 | -7.97 to -2.09 | -5.50 | 1.40 | <0.001 | -8.34 to -2.68 |
| SLPI§ | 6.40 | 1.20 | <0.001 | 3.97 to 8.81 | 6.30 | 1.20 | <0.001 | 3.87 to 8.75 |
| IL5 | 0.48 | 5.20 | 0.93 | -9.69 to 10.6 | 1.10 | 5.20 | 0.84 | -9.10 to 11.2 |
| S100A8 | 6.40 | 2.50 | 0.01 | 1.42 to 11.4 | 5.60 | 2.50 | 0.03 | 0.69 to 10.6 |
| TARC | 5.00 | 7.80 | 0.52 | -10.2 to 20.3 | 7.50 | 7.40 | 0.32 | -7.06 to 22.0 |
| GMCSF | 2.80 | 5.70 | 0.63 | -8.36 to 13.9 | 3.90 | 6.40 | 0.54 | -8.64 to 16.5 |
| HMGB1 | 0.12 | 3.00 | 0.97 | -5.68 to 5.91 | 0.77 | 2.90 | 0.79 | -5.01 to 6.55 |
| TNFα | 3.10 | 2.30 | 0.18 | -1.41 to 7.65 | 3.60 | 2.30 | 0.12 | -0.90 to 8.15 |
| IFNγ | 0.81 | 4.50 | 0.86 | -8.05 to 9.67 | 1.20 | 4.50 | 0.79 | -7.69 to 10.1 |
| IL6 | 3.50 | 2.60 | 0.19 | -1.66 to 8.6 | 3.40 | 2.60 | 0.20 | -1.77 to 8.52 |
| IL10 | 5.30 | 2.70 | 0.05 | 0.01 to 10.5 | 5.50 | 2.80 | 0.05 | -0.02 to 11.0 |
| CRP | -2.20 | 3.40 | 0.53 | -8.91 to 4.61 | -3.20 | 6.20 | 0.61 | -15.4 to 9.05 |

* **Abbreviations**: **CRP** C reactive protein; **ENRAGE** Extracellular newly identified receptor for advanced glycation end products binding protein; **GMCSF** granulocyte macrophage colony stimulating factor; **HMGB1** high mobility group box 1 protein; **ICAM1** intracellular adhesion molecule 1; **IFNγ** interferon γ; **IL** interleukin; **MMP9** matrix metallopeptidase 9; **MPO** myeloperoxidase; **NE** neutrophil elastase; **PR3** proteinase 3; **sRAGE** soluble receptor for advanced glycation products; **SLPI** secretory leukoprotease inhibitor; **TARC** thymus and activation regulation chemokine; **TNFα** tumor necrosis factor α; **YKL40** chitinase 3-like 1 protein. This table provides complete results using data adjusted for biomarker measurements outside the limits of detection as well as data adjusted and imputed for data missingness for main text Table 6, $FEV_1$%.

† Linear Regression.

§ Significant with FDR = 0.001.

¶ Significant with FDR = 0.01.
