## Supplemental Table 8 for "Associations of Sputum Biomarkers with Clinical Outcomes in People with Cystic Fibrosis"

**Supplemental Table 8A. Biomarkers as Univariable Adjustments to Weight-for-Age *z*-Score in Proportional Hazards Models of Time to Next Exacerbation.**[*]

| Univariable Biomarker[†] | Data Adjusted for Detection Limits | | | | | Data Adjusted and Imputed for Missingness | | | | |
|---|---|---|---|---|---|---|---|---|---|---|
| | Estimate | SE | Hazard Ratio | *p* | 95% CI of the Hazard Ratio | Estimate | SE | Hazard Ratio | *p* | 95% CI of the Hazard Ratio |
| MPO | 0.29 | 0.09 | 1.33 | 0.002 | 1.11 to 1.60 | 0.28 | 0.09 | 1.32 | 0.003 | 1.10 to 1.58 |
| NE | 0.17 | 0.08 | 1.18 | 0.025 | 1.02 to 1.37 | 0.17 | 0.08 | 1.18 | 0.025 | 1.02 to 1.37 |
| S100A9 | -0.27 | 0.12 | 0.76 | 0.028 | 0.60 to 0.97 | -0.19 | 0.13 | 0.83 | 0.16 | 0.64 to 1.08 |
| SRAGE | 0.19 | 0.09 | 1.21 | 0.037 | 1.01 to 1.44 | 0.20 | 0.09 | 1.22 | 0.025 | 1.03 to 1.46 |
| ENRAGE | 0.25 | 0.09 | 1.29 | 0.006 | 1.07 to 1.55 | 0.25 | 0.09 | 1.29 | 0.007 | 1.07 to 1.54 |
| IL1β | 0.17 | 0.08 | 1.18 | 0.029 | 1.02 to 1.37 | 0.17 | 0.08 | 1.18 | 0.028 | 1.02 to 1.37 |
| YKL40 | 0.19 | 0.09 | 1.20 | 0.048 | 1.00 to 1.45 | 0.21 | 0.09 | 1.24 | 0.02 | 1.03 to 1.48 |
| IL17A | 0.43 | 0.18 | 1.54 | 0.017 | 1.08 to 2.20 | 0.42 | 0.18 | 1.53 | 0.017 | 1.08 to 2.16 |
| PR3 | 0.12 | 0.09 | 1.13 | 0.16 | 0.95 to 1.33 | 0.17 | 0.10 | 1.18 | 0.076 | 0.98 to 1.43 |
| ICAM1 | 0.32 | 0.16 | 1.38 | 0.039 | 1.02 to 1.87 | 0.30 | 0.14 | 1.35 | 0.032 | 1.03 to 1.77 |
| IL8 | 0.42 | 0.24 | 1.52 | 0.077 | 0.96 to 2.41 | 0.41 | 0.23 | 1.51 | 0.079 | 0.95 to 2.38 |
| Calprotectin | 0.058 | 0.04 | 1.06 | 0.10 | 0.99 to 1.14 | 0.06 | 0.04 | 1.06 | 0.10 | 0.99 to 1.14 |
| MMP9 | 0.16 | 0.09 | 1.17 | 0.066 | 0.99 to 1.39 | 0.20 | 0.08 | 1.22 | 0.017 | 1.04 to 1.44 |
| SLPI | -0.10 | 0.08 | 0.91 | 0.25 | 0.77 to 1.07 | -0.10 | 0.08 | 0.91 | 0.25 | 0.77 to 1.07 |
| IL5 | 0.39 | 0.30 | 1.48 | 0.20 | 0.82 to 2.68 | 0.36 | 0.30 | 1.44 | 0.23 | 0.80 to 2.58 |
| S100A8 | 0.001 | 0.16 | 1.00 | 1.00 | 0.74 to 1.36 | 0.01 | 0.15 | 1.01 | 0.97 | 0.74 to 1.36 |
| TARC | 0.43 | 0.44 | 1.54 | 0.33 | 0.64 to 3.67 | 0.32 | 0.42 | 1.38 | 0.44 | 0.61 to 3.15 |
| GMCSF | 0.46 | 0.28 | 1.59 | 0.10 | 0.91 to 2.77 | 0.39 | 0.33 | 1.48 | 0.23 | 0.78 to 2.81 |
| HMGB1 | 0.10 | 0.16 | 1.11 | 0.51 | 0.82 to 1.51 | 0.10 | 0.15 | 1.10 | 0.52 | 0.82 to 1.49 |
| TNFα | -0.04 | 0.12 | 0.96 | 0.75 | 0.77 to 1.21 | -0.05 | 0.12 | 0.95 | 0.65 | 0.76 to 1.19 |
| IFNγ | 0.25 | 0.25 | 1.28 | 0.33 | 0.78 to 2.09 | 0.22 | 0.25 | 1.25 | 0.37 | 0.77 to 2.04 |
| IL6 | 0.02 | 0.14 | 1.02 | 0.87 | 0.78 to 1.35 | 0.02 | 0.14 | 1.02 | 0.89 | 0.77 to 1.34 |
| IL10 | 0.09 | 0.16 | 1.09 | 0.59 | 0.80 to 1.49 | 0.08 | 0.16 | 1.08 | 0.63 | 0.79 to 1.48 |
| CRP | 0.01 | 0.18 | 1.01 | 0.95 | 0.71 to 1.44 | -0.02 | 0.34 | 0.98 | 0.96 | 0.50 to 1.92 |

[*] This table provides complete results using data adjusted for biomarker measurements outside the limits of detection as well as data adjusted and imputed for data missingness.

[†] **Abbreviations**: **CRP** C reactive protein; **ENRAGE** Extracellular newly identified receptor for advanced glycation end products binding protein; **GMCSF** granulocyte macrophage colony stimulating factor; **HMGB1** high mobility group box 1 protein; **ICAM1** intracellular adhesion molecule 1; **IFNγ** interferon γ; **IL** interleukin; **MMP9** matrix metallopeptidase 9; **MPO** myeloperoxidase; **NE** neutrophil elastase; **PR3** proteinase 3; **sRAGE** soluble receptor for advanced glycation products; **SLPI** secretory leukoprotease inhibitor; **TARC** thymus and activation regulation chemokine; **TNFα** tumor necrosis factor α; **YKL40** chitinase 3-like 1 protein.

**Supplemental Table 8B. Biomarkers as Univariable Adjustments to Diabetes in Proportional Hazards Models of Time to Next Exacerbation.**[*]

| Univariable Biomarker | Data Adjusted for Detection Limits | | | | | Data Adjusted and Imputed for Missingness | | | | |
|---|---|---|---|---|---|---|---|---|---|---|
| | **Estimate** | **SE** | **Hazard Ratio** | ***p*** | **95% CI of the Hazard Ratio** | **Estimate** | **SE** | **Hazard Ratio** | ***p*** | **95% CI of the Hazard Ratio** |
| MPO | 0.22 | 0.10 | 1.24 | 0.027 | 1.03 to 1.50 | 0.21 | 0.10 | 1.23 | 0.028 | 1.02 to 1.48 |
| NE | 0.17 | 0.08 | 1.19 | 0.027 | 1.02 to 1.38 | 0.17 | 0.08 | 1.19 | 0.027 | 1.02 to 1.38 |
| S100A9 | -0.25 | 0.13 | 0.78 | 0.046 | 0.61 to 1.00 | -0.16 | 0.13 | 0.85 | 0.220 | 0.66 to 1.10 |
| SRAGE | 0.19 | 0.09 | 1.21 | 0.033 | 1.02 to 1.45 | 0.21 | 0.09 | 1.23 | 0.022 | 1.03 to 1.47 |
| ENRAGE | 0.18 | 0.09 | 1.19 | 0.059 | 0.99 to 1.43 | 0.18 | 0.09 | 1.19 | 0.057 | 0.99 to 1.43 |
| IL1β | 0.15 | 0.08 | 1.17 | 0.052 | 1.00 to 1.36 | 0.15 | 0.08 | 1.17 | 0.051 | 1.00 to 1.36 |
| YKL40 | 0.17 | 0.10 | 1.19 | 0.087 | 0.98 to 1.44 | 0.18 | 0.10 | 1.19 | 0.071 | 0.98 to 1.45 |
| IL17A | 0.30 | 0.16 | 1.34 | 0.073 | 0.97 to 1.86 | 0.26 | 0.16 | 1.30 | 0.110 | 0.95 to 1.77 |
| PR3 | 0.12 | 0.09 | 1.13 | 0.17 | 0.95 to 1.35 | 0.15 | 0.10 | 1.16 | 0.150 | 0.95 to 1.41 |
| ICAM1 | 0.24 | 0.16 | 1.27 | 0.12 | 0.94 to 1.72 | 0.21 | 0.14 | 1.24 | 0.120 | 0.94 to 1.63 |
| IL8 | 0.28 | 0.23 | 1.32 | 0.23 | 0.84 to 2.09 | 0.26 | 0.23 | 1.30 | 0.250 | 0.83 to 2.04 |
| Calprotectin | 0.06 | 0.04 | 1.06 | 0.11 | 0.99 to 1.14 | 0.06 | 0.04 | 1.06 | 0.110 | 0.99 to 1.14 |
| MMP9 | 0.11 | 0.08 | 1.12 | 0.19 | 0.95 to 1.32 | 0.14 | 0.08 | 1.15 | 0.085 | 0.98 to 1.36 |
| SLPI | -0.07 | 0.08 | 0.93 | 0.37 | 0.79 to 1.09 | -0.08 | 0.08 | 0.93 | 0.330 | 0.79 to 1.08 |
| IL5 | 0.35 | 0.30 | 1.41 | 0.25 | 0.78 to 2.55 | 0.33 | 0.30 | 1.39 | 0.270 | 0.77 to 2.51 |
| S100A8 | -0.16 | 0.14 | 0.86 | 0.27 | 0.65 to 1.13 | -0.16 | 0.14 | 0.86 | 0.250 | 0.65 to 1.12 |
| TARC | 0.43 | 0.45 | 1.54 | 0.33 | 0.64 to 3.72 | 0.44 | 0.42 | 1.55 | 0.290 | 0.69 to 3.5 |
| GMCSF | 0.19 | 0.27 | 1.21 | 0.47 | 0.72 to 2.03 | 0.08 | 0.30 | 1.08 | 0.790 | 0.60 to 1.97 |
| HMGB1 | 0.10 | 0.15 | 1.10 | 0.53 | 0.82 to 1.48 | 0.09 | 0.15 | 1.09 | 0.550 | 0.81 to 1.47 |
| TNFα | -0.07 | 0.12 | 0.93 | 0.56 | 0.74 to 1.17 | -0.07 | 0.12 | 0.93 | 0.540 | 0.74 to 1.17 |
| IFNγ | 0.19 | 0.25 | 1.21 | 0.45 | 0.74 to 1.97 | 0.18 | 0.25 | 1.19 | 0.480 | 0.73 to 1.94 |
| IL6 | -0.02 | 0.14 | 0.98 | 0.89 | 0.74 to 1.29 | 0.00 | 0.14 | 1.00 | 0.970 | 0.76 to 1.31 |
| IL10 | 0.06 | 0.15 | 1.06 | 0.70 | 0.79 to 1.43 | 0.07 | 0.16 | 1.07 | 0.680 | 0.79 to 1.45 |
| CRP | 0.00 | 0.18 | 1.00 | 0.99 | 0.70 to 1.43 | 0.02 | 0.34 | 1.02 | 0.960 | 0.52 to 1.99 |

[*] This table provides complete results using data adjusted for biomarker measurements outside the limits of detection as well as data adjusted and imputed for data missingness.

[†] **Abbreviations**: **CRP** C reactive protein; **ENRAGE** Extracellular newly identified receptor for advanced glycation end products binding protein; **GMCSF** granulocyte macrophage colony stimulating factor; **HMGB1** high mobility group box 1 protein; **ICAM1** intracellular adhesion molecule 1; **IFNγ** interferon γ; **IL** interleukin; **MMP9** matrix metallopeptidase 9; **MPO** myeloperoxidase; **NE** neutrophil elastase; **PR3** proteinase 3; **sRAGE** soluble receptor for advanced glycation products; **SLPI** secretory leukoprotease inhibitor; **TARC** thymus and activation regulation chemokine; **TNFα** tumor necrosis factor α; **YKL40** chitinase 3-like 1 protein.

**Supplemental Table 8C. Biomarkers as Univariable Adjustments to $FEV_1$% in Proportional Hazards Models of Time to Next Exacerbation.***

| Univariable Biomarker† | Data Adjusted for Detection Limits | | | | | Data Adjusted and Imputed for Missingness | | | | |
|---|---|---|---|---|---|---|---|---|---|---|
| | Estimate | SE | Hazard Ratio | *p* | 95% CI of the Hazard Ratio | Estimate | SE | Hazard Ratio | *p* | 95% CI of the Hazard Ratio |
| MPO | 0.19 | 0.10 | 1.21 | 0.06 | 0.99 to 1.48 | 0.18 | 0.10 | 1.20 | 0.07 | 0.98 to 1.46 |
| NE | 0.14 | 0.08 | 1.15 | 0.09 | 0.98 to 1.35 | 0.14 | 0.08 | 1.15 | 0.09 | 0.98 to 1.35 |
| S100A9 | -0.24 | 0.12 | 0.79 | 0.051 | 0.62 to 1.00 | -0.18 | 0.12 | 0.83 | 0.14 | 0.65 to 1.06 |
| SRAGE | 0.14 | 0.10 | 1.15 | 0.14 | 0.95 to 1.39 | 0.15 | 0.10 | 1.16 | 0.12 | 0.96 to 1.40 |
| ENRAGE | 0.15 | 0.10 | 1.16 | 0.12 | 0.96 to 1.41 | 0.15 | 0.10 | 1.16 | 0.12 | 0.96 to 1.40 |
| IL1β | 0.13 | 0.08 | 1.14 | 0.11 | 0.97 to 1.33 | 0.13 | 0.08 | 1.14 | 0.11 | 0.97 to 1.33 |
| YKL40 | 0.15 | 0.1 | 1.16 | 0.14 | 0.95 to 1.41 | 0.16 | 0.10 | 1.18 | 0.09 | 0.97 to 1.43 |
| IL17A | 0.24 | 0.17 | 1.28 | 0.16 | 0.91 to 1.79 | 0.22 | 0.17 | 1.25 | 0.19 | 0.90 to 1.74 |
| PR3 | 0.11 | 0.09 | 1.12 | 0.22 | 0.94 to 1.35 | 0.11 | 0.10 | 1.12 | 0.27 | 0.92 to 1.37 |
| ICAM1 | 0.18 | 0.15 | 1.20 | 0.23 | 0.89 to 1.63 | 0.17 | 0.14 | 1.19 | 0.22 | 0.90 to 1.56 |
| IL8 | 0.29 | 0.24 | 1.33 | 0.23 | 0.83 to 2.14 | 0.27 | 0.24 | 1.31 | 0.25 | 0.82 to 2.09 |
| Calprotectin | 0.04 | 0.04 | 1.04 | 0.34 | 0.96 to 1.11 | 0.04 | 0.04 | 1.04 | 0.34 | 0.96 to 1.11 |
| MMP9 | 0.08 | 0.09 | 1.08 | 0.37 | 0.91 to 1.29 | 0.11 | 0.09 | 1.11 | 0.22 | 0.94 to 1.33 |
| SLPI | -0.03 | 0.09 | 0.98 | 0.77 | 0.82 to 1.15 | -0.02 | 0.09 | 0.98 | 0.79 | 0.83 to 1.15 |
| IL5 | 0.25 | 0.31 | 1.28 | 0.43 | 0.70 to 2.35 | 0.22 | 0.31 | 1.24 | 0.48 | 0.68 to 2.27 |
| S100A8 | -0.11 | 0.15 | 0.89 | 0.44 | 0.67 to 1.19 | -0.10 | 0.14 | 0.91 | 0.50 | 0.68 to 1.20 |
| TARC | 0.31 | 0.45 | 1.37 | 0.48 | 0.57 to 3.27 | 0.26 | 0.42 | 1.30 | 0.53 | 0.58 to 2.94 |
| GMCSF | 0.24 | 0.28 | 1.27 | 0.40 | 0.73 to 2.2 | 0.15 | 0.33 | 1.16 | 0.65 | 0.61 to 2.19 |
| HMGB1 | 0.08 | 0.16 | 1.09 | 0.60 | 0.80 to 1.47 | 0.10 | 0.15 | 1.10 | 0.52 | 0.82 to 1.48 |
| TNFα | -0.09 | 0.12 | 0.91 | 0.46 | 0.72 to 1.16 | -0.10 | 0.12 | 0.91 | 0.41 | 0.71 to 1.15 |
| IFNγ | 0.06 | 0.26 | 1.06 | 0.83 | 0.63 to 1.77 | 0.03 | 0.26 | 1.03 | 0.90 | 0.621-1.72 |
| IL6 | -0.08 | 0.14 | 0.93 | 0.59 | 0.70 to 1.23 | -0.07 | 0.14 | 0.93 | 0.63 | 0.708-1.23 |
| IL10 | -0.04 | 0.15 | 0.96 | 0.77 | 0.71 to 1.29 | -0.05 | 0.16 | 0.96 | 0.77 | 0.705-1.29 |
| CRP | 0.00 | 0.18 | 1.00 | 0.99 | 0.70 to 1.44 | 0.07 | 0.35 | 1.07 | 0.84 | 0.543-2.12 |

[*] This table provides complete results using data adjusted for biomarker measurements outside the limits of detection as well as data adjusted and imputed for data missingness.

[†] **Abbreviations**: **CRP** C reactive protein; **ENRAGE** Extracellular newly identified receptor for advanced glycation end products binding protein; **GMCSF** granulocyte macrophage colony stimulating factor; **HMGB1** high mobility group box 1 protein; **ICAM1** intracellular adhesion molecule 1; **IFNγ** interferon γ; **IL** interleukin; **MMP9** matrix metallopeptidase 9; **MPO** myeloperoxidase; **NE** neutrophil elastase; **PR3** proteinase 3; **sRAGE** soluble receptor for advanced glycation products; **SLPI** secretory leukoprotease inhibitor; **TARC** thymus and activation regulation chemokine; **TNFα** tumor necrosis factor α; **YKL40** chitinase 3-like 1 protein.
