## Supplemental Table 9 for "Associations of Sputum Biomarkers with Clinical Outcomes in People with Cystic Fibrosis"

**Supplemental Table 9. Biomarkers in Adjusted* Proportional Hazards Models of Time to Next Exacerbation***

| Univariable Biomarker† | Data Adjusted for Detection Limits | | | | | Data Adjusted and Imputed for Missingness | | | | |
|---|---|---|---|---|---|---|---|---|---|---|
| | Estimate | SE | Hazard Ratio | *p* | 95% CI of the Hazard Ratio | Estimate | SE | Hazard Ratio | *p* | 95% CI of the Hazard Ratio |
| ENRAGE | 0.30 | 0.10 | 1.35 | 0.003 | 1.10 to 1.65 | 0.30 | 0.10 | 1.34 | 0.004 | 1.10 to 1.64 |
| MPO | 0.26 | 0.10 | 1.30 | 0.008 | 1.07 to 1.57 | 0.28 | 0.10 | 1.32 | 0.006 | 1.08 to 1.61 |
| SRAGE | 0.20 | 0.09 | 1.23 | 0.021 | 1.03 to 1.46 | 0.25 | 0.10 | 1.29 | 0.009 | 1.06 to 1.55 |
| ICAM1 | 0.38 | 0.16 | 1.46 | 0.021 | 1.06 to 2.02 | 0.22 | 0.09 | 1.24 | 0.014 | 1.04 to 1.47 |
| NE | 0.18 | 0.08 | 1.19 | 0.022 | 1.03 to 1.39 | 0.21 | 0.09 | 1.24 | 0.015 | 1.04 to 1.47 |
| YKL40 | 0.23 | 0.10 | 1.26 | 0.024 | 1.03 to 1.53 | 0.34 | 0.15 | 1.41 | 0.020 | 1.06 to 1.88 |
| TARC | 1.00 | 0.46 | 2.72 | 0.031 | 1.10 to 6.74 | 0.18 | 0.08 | 1.19 | 0.022 | 1.03 to 1.39 |
| MMP9 | 0.18 | 0.09 | 1.20 | 0.039 | 1.01 to 1.43 | 0.16 | 0.08 | 1.17 | 0.047 | 1.00 to 1.37 |
| IL1β | 0.16 | 0.08 | 1.17 | 0.049 | 1.00 to 1.37 | 0.82 | 0.44 | 2.27 | 0.062 | 0.96 to 5.38 |
| IL5 | 0.58 | 0.31 | 1.79 | 0.062 | 0.97 to 3.29 | 0.54 | 0.31 | 1.72 | 0.077 | 0.94 to 3.15 |
| IL17A | 0.28 | 0.17 | 1.32 | 0.098 | 0.95 to 1.83 | 0.28 | 0.16 | 1.32 | 0.091 | 0.96 to 1.82 |
| S100A9 | -0.20 | 0.12 | 0.82 | 0.12 | 0.645 to 1.05 | 0.17 | 0.10 | 1.19 | 0.092 | 0.97 to 1.45 |
| IFNγ | 0.37 | 0.26 | 1.44 | 0.16 | 0.861 to 2.41 | 0.34 | 0.26 | 1.40 | 0.19 | 0.84 to 2.34 |
| PR3 | 0.12 | 0.09 | 1.13 | 0.19 | 0.94 to 1.36 | 0.29 | 0.25 | 1.34 | 0.24 | 0.82 to 2.19 |
| IL8 | 0.30 | 0.25 | 1.36 | 0.23 | 0.823 to 2.23 | -0.09 | 0.09 | 0.92 | 0.33 | 0.77 to 1.09 |
| GMCSF | 0.29 | 0.27 | 1.34 | 0.28 | 0.784 to 2.30 | 0.03 | 0.04 | 1.03 | 0.34 | 0.97 to 1.11 |
| SLPI | -0.09 | 0.09 | 0.91 | 0.30 | 0.762 to 1.09 | -0.11 | 0.14 | 0.90 | 0.43 | 0.69 to 1.17 |
| Calprotectin | 0.03 | 0.04 | 1.03 | 0.34 | 0.966 to 1.11 | -0.11 | 0.17 | 0.90 | 0.53 | 0.65 to 1.25 |
| IL10 | 0.09 | 0.15 | 1.10 | 0.54 | 0.817 to 1.47 | 0.19 | 0.32 | 1.21 | 0.55 | 0.64 to 2.29 |
| S100A8 | -0.06 | 0.14 | 0.94 | 0.67 | 0.709 to 1.25 | 0.09 | 0.15 | 1.09 | 0.58 | 0.80 to 1.47 |
| TNFα | -0.04 | 0.12 | 0.96 | 0.74 | 0.765 to 1.21 | -0.05 | 0.12 | 0.95 | 0.66 | 0.76 to 1.19 |
| HMGB1 | -0.06 | 0.17 | 0.95 | 0.75 | 0.677 to 1.32 | -0.04 | 0.14 | 0.96 | 0.78 | 0.73 to 1.27 |
| IL6 | 0.04 | 0.14 | 1.04 | 0.81 | 0.78 to 1.37 | 0.04 | 0.14 | 1.04 | 0.8 | 0.78 to 1.37 |
| CRP | -0.04 | 0.19 | 0.96 | 0.84 | 0.662 to 1.40 | 0.06 | 0.36 | 1.07 | 0.86 | 0.53 to 2.15 |

* Adjusted by Age, Sex and Number of Prior Pulmonary Exacerbations.

† **Abbreviations**: **CRP** C reactive protein; **ENRAGE** Extracellular newly identified receptor for advanced glycation end products binding protein; **GMCSF** granulocyte macrophage colony stimulating factor; **HMGB1** high mobility group box 1 protein; **ICAM1** intracellular adhesion molecule 1; **IFNγ** interferon γ; **IL** interleukin; **MMP9** matrix metallopeptidase 9; **MPO** myeloperoxidase; **NE** neutrophil elastase; **PR3** proteinase 3; **sRAGE** soluble receptor for advanced glycation products; **SLPI** secretory leukoprotease inhibitor; **TARC** thymus and activation regulation chemokine; **TNFα** tumor necrosis factor α; **YKL40** chitinase 3-like 1 protein.
