## Supplemental Figure 1 for "Associations of Sputum Biomarkers with Clinical Outcomes in People with Cystic Fibrosis"

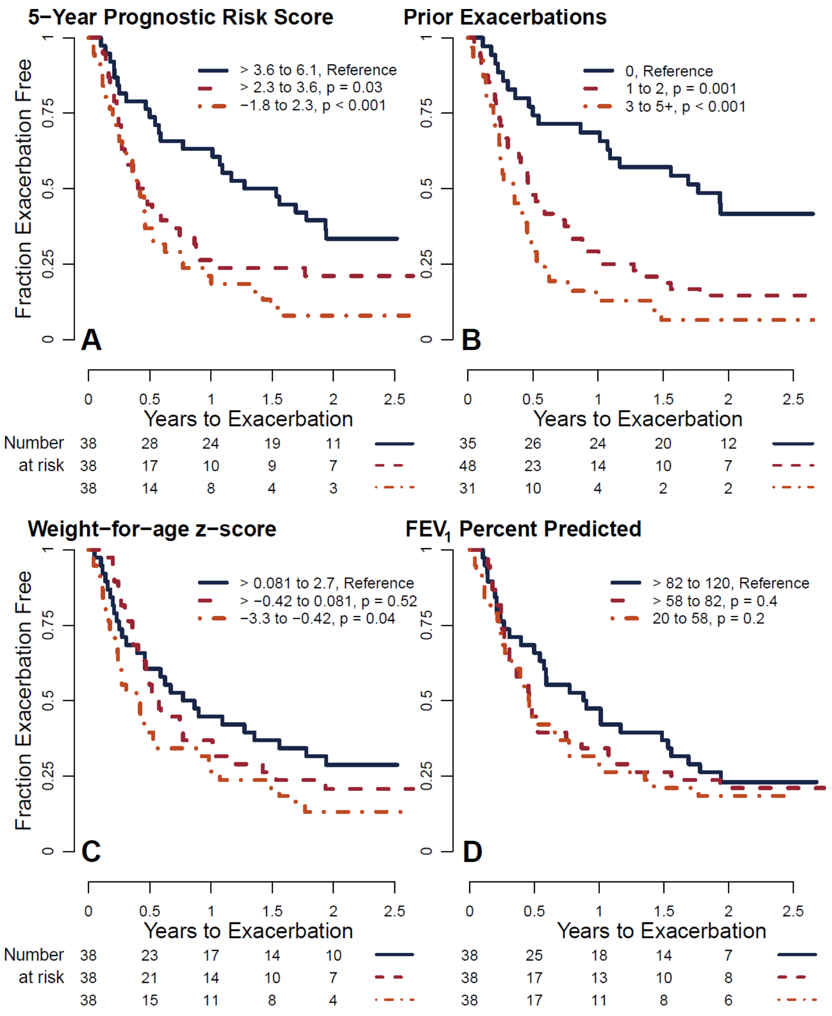


**Supplemental Figure 1. Kaplan-Meier Plots Exploring Relationships with Time to Next Pulmonary Exacerbation.** Patients were stratified into evenly sized groups as shown in each panel legend for **(A)** 5-Year Prognostic Risk Score, **(B)** Number of Pulmonary Exacerbations in the year prior to enrollment, **(C)** Weight-for-age *z*-score and **(D)** $FEV_1$%. Groups are equal in size except for number of prior exacerbations because those values are ordinal and do not allow more even distribution.
