## Supplemental Figure 2 for "Associations of Sputum Biomarkers with Clinical Outcomes in People with Cystic Fibrosis"

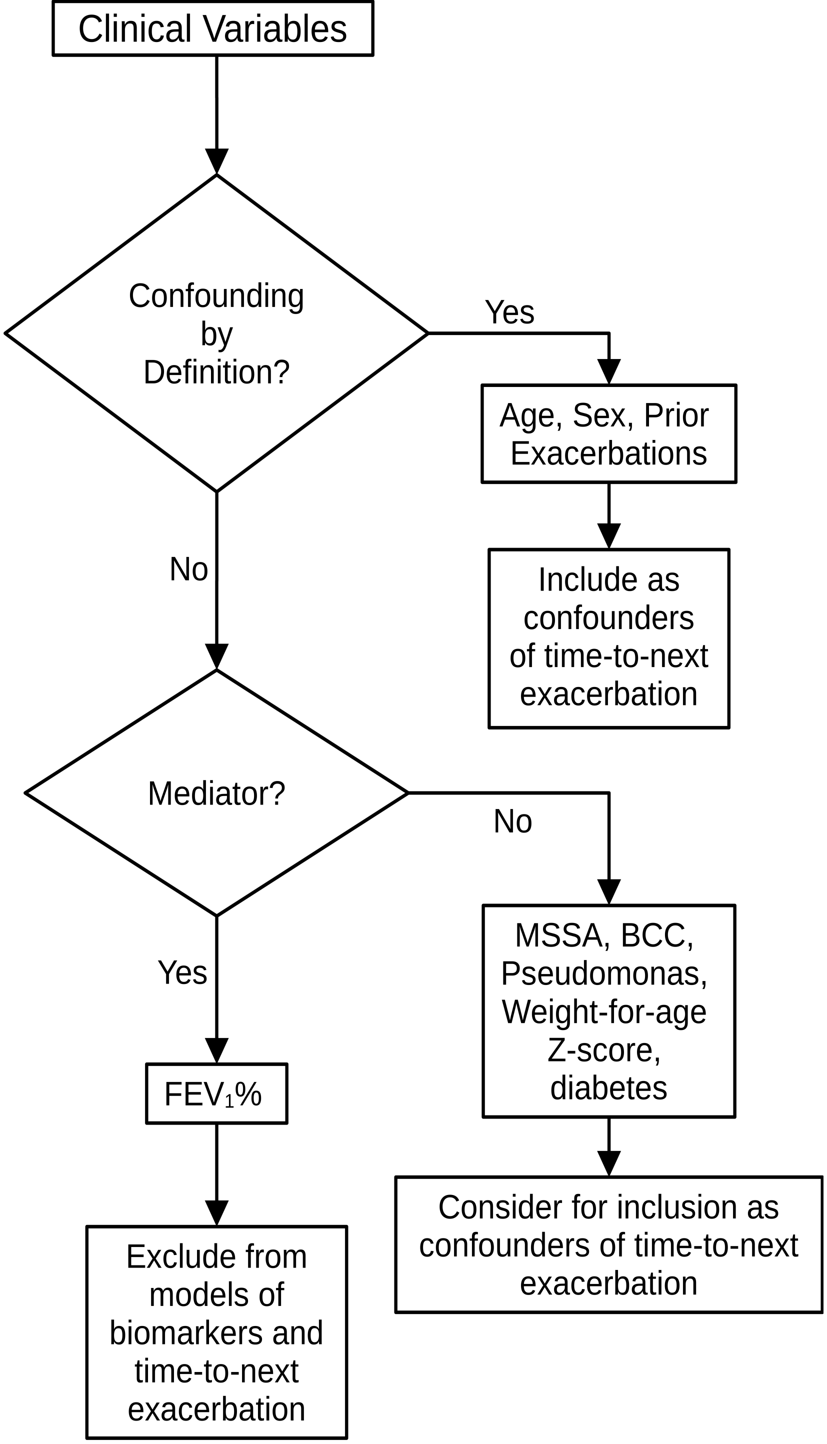


**Supplemental Figure 2. Confounding and Mediation Analysis.** Clinical variables were evaluated for their role as confounders and mediators of the association of potential biomarkers of inflammation and time-to-next pulmonary exacerbation. See the website: Mediation (David A. Kenny) [Internet]https://davidakenny.net/cm/mediate.htm cited March 14, 2022 and Baron RM, Kenny DA. The moderator-mediator variable distinction in social psychological research: conceptual, strategic, and statistical considerations. *J Pers Soc Psychol* 1986;51(6):1173–1182 for the adapted procedures.
