## Supplemental Figure 3 for "Associations of Sputum Biomarkers with Clinical Outcomes in People with Cystic Fibrosis"

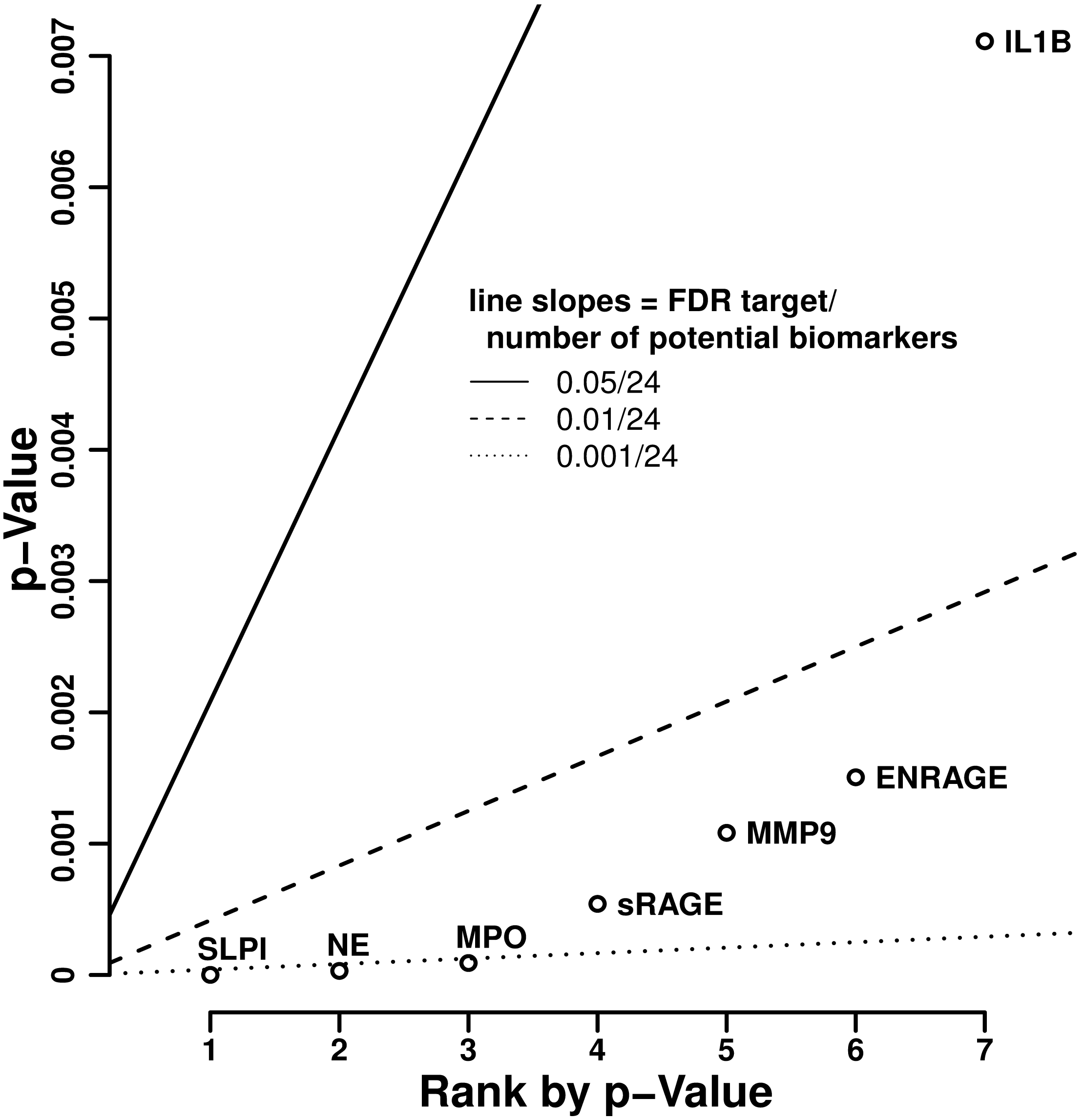


**Supplemental Figure 3. False Discovery Rate Analysis of Potential Biomarkers for Associations with $FEV_1$%** Using the *p*-values of each inflammatory marker from univariable linear regression models for $FEV_1$%, we ranked the potential biomarkers by *p*-value and drew lines with slopes determined by the threshhold for false discovery (set to 0.05, 0.01 and 0.001) divided by the number of potential biomarkers in the entire study. Biomarkers falling below a line have chance of being true findings that equal 1 – false discovery set point for that line. Three biomarkers fall below the FDR 0.001 line, three fall below FDR 0.01 and one falls below the FDR 0.05 line in this case.
