## Supplemental Figure 4 for "Associations of Sputum Biomarkers with Clinical Outcomes in People with Cystic Fibrosis"

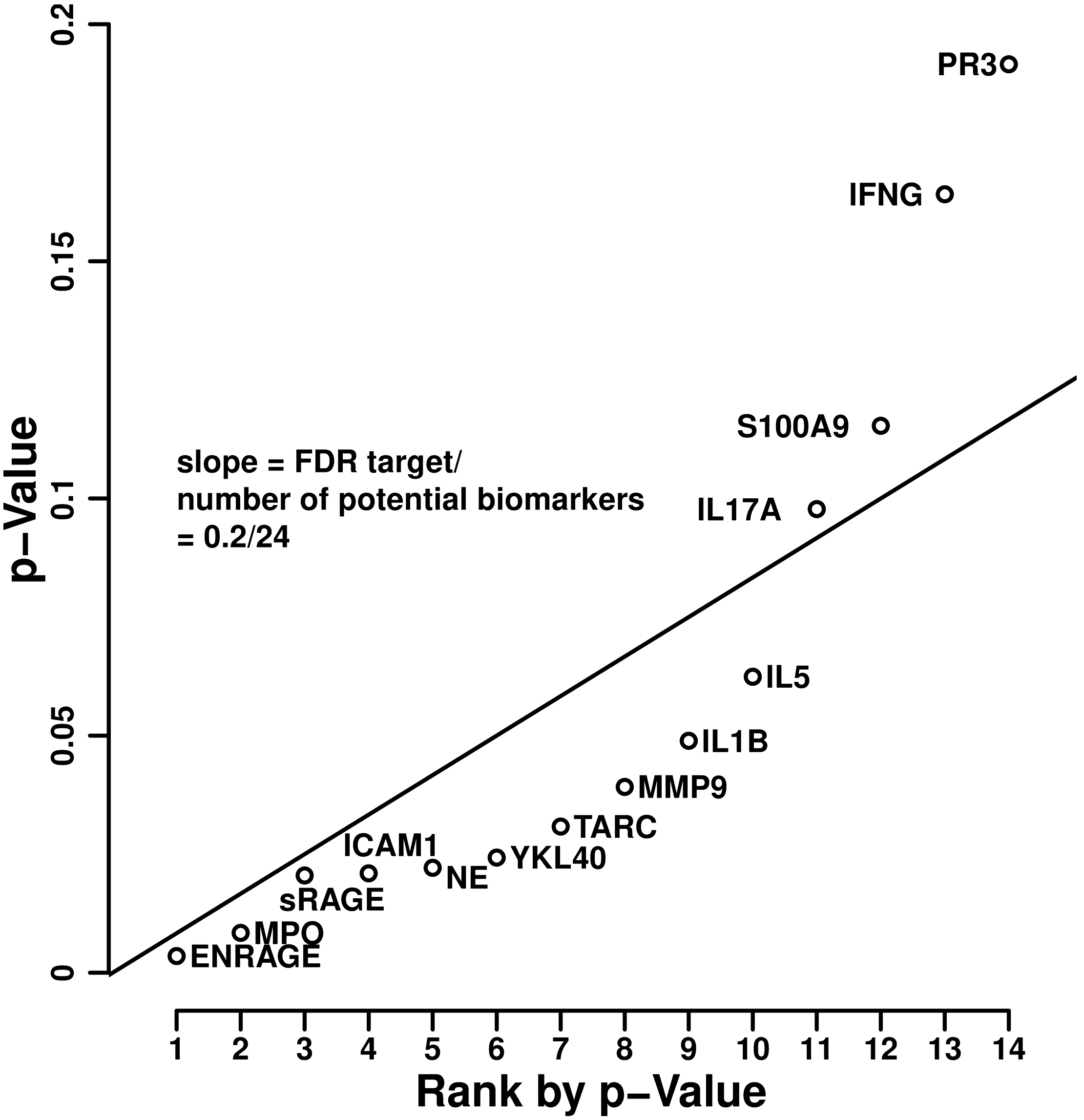


**Supplemental Figure 4. False Discovery Rate Analysis of Potential Biomarkers for Associations with Time to Next Exacerbation** Using the *p*-values of each inflammatory marker from proportional hazards models for time to next exacerbation with adjustments for age, sex and number of prior exacerbations in the year prior to study enrollment, we ranked the potential biomarkers by *p*-value and drew a line with a slope determined by the threshhold for false discovery (set to 0.2) divided by the number of potential biomarkers in the entire study. Any biomarker falling below the line has a chance of being a true finding that is equal to 1 – false discovery set point. In this case, ten biomarkers fall below the line suggesting that 8 of them are likely to be true findings.
