## Supplemental Figure 5 for "Associations of Sputum Biomarkers with Clinical Outcomes in People with Cystic Fibrosis"

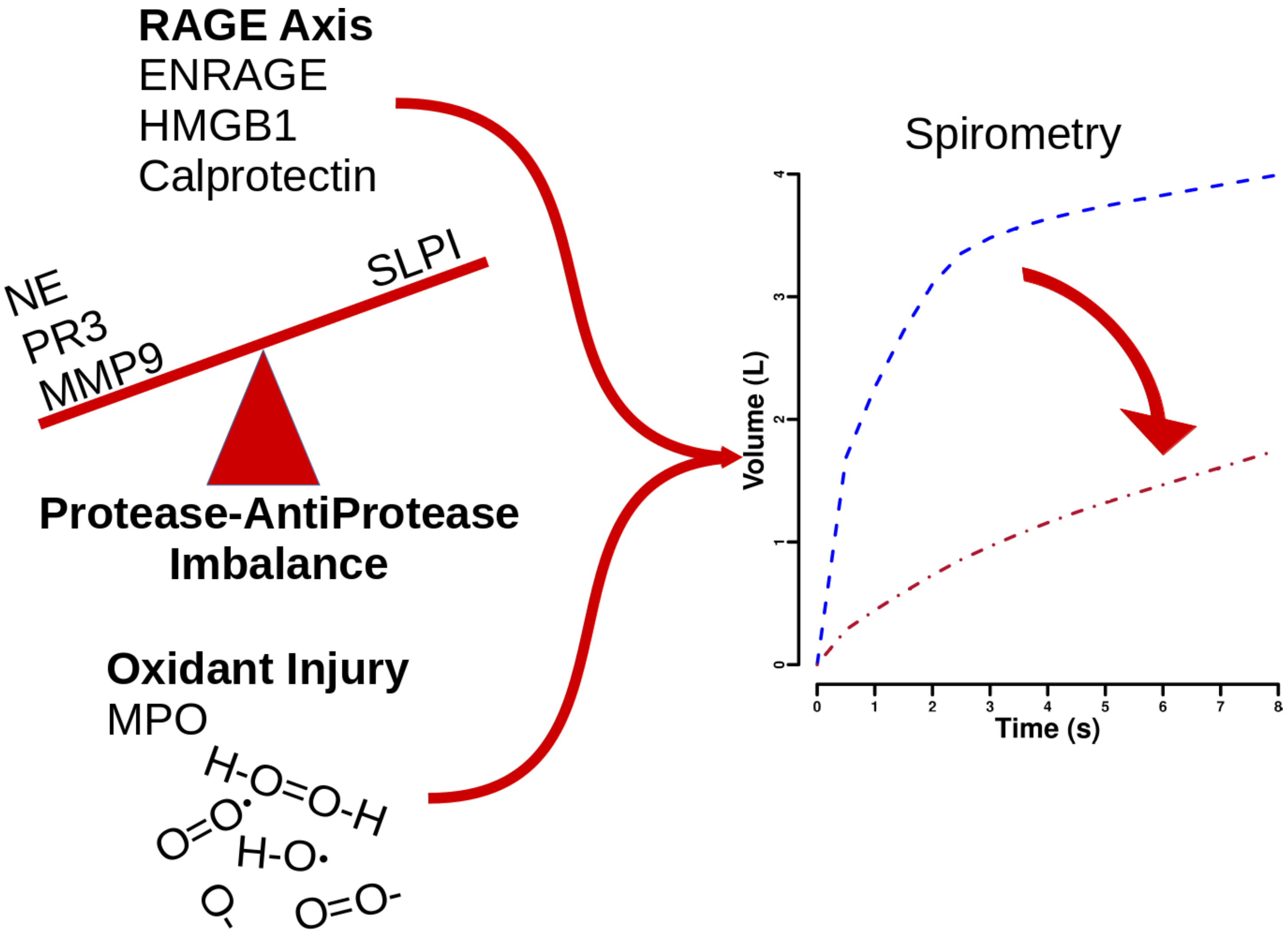


**Supplemental Figure 5. Relationship between RAGE Axis, Protease-Antiprotease Imbalance, Oxidant Injury and Lung Function.** Three pathways of injury with representative biomarkers shown contribute to reducing lung function.
